# Optimal deployment of gonorrhoea point-of-care tests: modelling the potential impact of diagnostic confirmation testing and screening strategies across five priority populations in Kenya

**DOI:** 10.64898/2025.12.05.25341711

**Authors:** Julia Michalow, Anne Cori, Joshua Kimani, Parinita Bhattacharjee, Marie-Claude Boily, Jeffrey W. Imai-Eaton

## Abstract

**Background:** Gonorrhoea treatment in sub-Saharan Africa relies on syndromic management, which has poor diagnostic performance and misses asymptomatic infections. Point-of-care tests (POCTs) could address these limitations, but anticipated supply constraints necessitate strategic allocation to maximise impact.

**Methods:** We developed a deterministic compartmental model of gonorrhoea transmission in Kenya to evaluate allocating POCTs for diagnostic confirmation of symptomatic care attendees versus screening of routine healthcare service attendees across five priority populations: female sex workers (FSW), their male clients (CFSW), pregnant women, adolescent girls and young women, or total population men. We modelled constrained and unrestricted POCT availability during 2025-2030, and estimated infections averted relative to baseline syndromic management. Quality-adjusted life years (QALYs) gained were quantified using probability-tree models.

**Results:** At baseline, incidence was highest among FSW (11.9 [UI:5.7-18.6] per 100 per year) and CFSW (13.1 [6.9-24.8]), while most QALY losses (80.6% [76.1-83.8%]) were among pregnant women and their infants. With constrained POCTs (sufficient to test 0.1% of adults annually), diagnostic confirmation averted the most transmission when among symptomatic FSW (2.1% [0.6-5.6%] of infections) or CFSW (2.2% [0.8-5.3%]), but the most morbidity was averted when among symptomatic pregnant women (3.5% [1.8-7.2%] of QALY losses). Screening averted <1% of infections or QALY losses across populations. With unrestricted POCTs, screening had larger absolute impacts but lower per-test returns than diagnostic confirmation.

**Conclusions:** Diagnostic confirmation should be prioritised over screening, supporting WHO guidance to strengthen aetiologic diagnosis within syndromic management. Prioritising diagnostic testing among symptomatic pregnant women had the largest impact on mitigating gonorrhoea-related morbidity.

## Background

The World Health Organization (WHO) classifies *Neisseria gonorrhoeae* as a high-priority pathogen due to its global burden and rapid emergence of antimicrobial resistance.^1^ Gonorrhoea disproportionately affects women, causing reproductive-tract complications such as pelvic inflammatory disease and infertility, and, when acquired during pregnancy, stillbirth and adverse neonatal outcomes.^2,3^ Prevalence in sub-Saharan Africa (SSA) is more than double the global average.^4,5^

Syndromic management, involving presumptive treatment based on presenting symptoms, remains the primary approach for treating symptomatic sexually transmitted infections (STIs) in SSA. The approach has poor sensitivity and specificity for cervical infections such as gonorrhoea, leaving many women inadequately treated.^6^ WHO therefore recommends integrating molecular diagnostics into syndromic algorithms^6^ and recently advised targeted gonorrhoea screening of priority populations in high-prevalence settings, given most infections are asymptomatic.^7^ However, neither strategy has been implemented at scale in SSA.^8^

Point-of-care tests (POCTs) could expand access to diagnostic testing and screening. Existing gonorrhoea POCTs fall short of WHO target product profile (TPP) criteria for cost, turnaround time, and accessibility, but new platforms are in development.^9–11^ Strategic allocation of new POCTs will be essential given likely supply and resource constraints.^9,10,12^ Key trade-offs include: prioritising incidence reduction by focusing testing among populations driving transmission versus morbidity reduction by testing populations at greatest risk of adverse sequelae; prioritising diagnostic confirmation among symptomatic individuals, who would have higher positivity rates versus screening asymptomatic individuals, who would otherwise remain untreated; and within these strategies, prioritising men, who are more likely to experience symptoms^13^ versus women, who are often inadequately managed under syndromic care due to poor diagnostic performance and face greater risks of reproductive and pregnancy-related sequelae.^6^ Previous modelling studies in SSA have evaluated diagnostic testing^14,15^ or screening,^16,17^ but none have directly compared these strategies or considered constrained POCT availability.

We developed an epidemic transmission dynamic model to evaluate allocation of constrained gonorrhoea POCTs across testing strategies and priority populations to maximise reductions in incidence and morbidity in Kenya between 2025 and 2030. We compared diagnostic confirmation of symptomatic individuals and screening of routine healthcare service attendees across five populations: female sex workers (FSW), their male clients (CFSW), pregnant women, sexually active adolescent girls and young women (AGYW), and men in the total population.

## Methods

### Compartmental model structure

We developed a deterministic compartmental model of heterosexual gonorrhoea transmission among 15-49-year-olds in Kenya (Text S1.1.1, Figure S1). The model represented an open, growing population stratified by sex (male, female), sexual risk (low, intermediate, high [FSW or CFSW]), age (15-24 years, 25-49 years), and gestational status (non-pregnant, pregnant). Individuals entered the model at 15-24 years as not yet sexually active, transitioning to sexual activity upon debut or when ageing to 25-49 years. Females transitioned between gestational states based on fertility rates. Individuals exited through all-cause mortality or ageing out at 50 years. Individuals did not transition between sexual risk groups.

Sexual partner acquisition rates were heterogenous across the population. We modelled proportionate mixing across all strata based on these rates, except FSW partnered exclusively with CFSW, while CFSW partnered with females in any risk group (Text S1.1.2). Same-sex partnerships were not modelled. The force of infection incorporated mixing patterns, sex acts per partnership, condom use, per-act transmission probability, and infection prevalence among partners (Text S1.1.3). After infection, individuals were either asymptomatic or symptomatic. Symptomatic individuals had a fixed probability of accessing care. Infections resolved rapidly with treatment or more slowly through natural clearance. We assumed no immunity and did not distinguish between anatomical sites of infection.

We modelled three diagnostic and treatment scenarios (Text S1.1.4, Figure S1). In the baseline scenario, syndromic management was provided to individuals accessing care for vaginal or urethral discharge. A proportion were appropriately treated according to the algorithm’s sensitivity. No treatment was provided to symptomatic individuals who did not access care or to asymptomatic individuals. In the diagnostic confirmation scenario, POCTs replaced syndromic management for a proportion of symptomatic care attendees. Those tested were diagnosed and treated according to test sensitivity, while those not tested were managed syndromically. Available tests were distributed across all symptomatic cases regardless of aetiology, with annual eligibility estimated using the gonorrhoea aetiologic proportion (proportion of discharge cases caused by gonorrhoea). In the screening scenario, POCTs were offered to routine primary healthcare or antenatal care attendees, with tests allocated according to population-specific attendance probabilities. Screen-positive individuals were treated. Syndromic management continued for symptomatic care attendees.

### Model parametrisation and calibration

The model incorporated 23 parameter types, which could vary by sex, risk, age, and/or gestational status. Of these, nine were fixed and fourteen groups were calibrated, yielding 41 total calibrated parameters. We specified uniform prior distributions for calibrated parameters, utilising multiple sources (Text S1.2). Demographic parameters were from UN World Population Prospects (WPP) 2024 projections (Text S1.2.1, Table S1).^18^ Sexual behaviour, mixing, and care-seeking parameters were from Kenya Demographic and Health Surveys (DHS; 2003, 2008–09, 2014, 2022)^19–22^ and the Kenya Population-based HIV Impact Assessment (2018) for the general population,^23^ and from published studies for FSW and CFSW (Text S1.2.2, Table S2). Natural history parameters were from literature (Table S2).

We calibrated the model to four equilibrium outcomes for the baseline scenario: (i) gonorrhoea prevalence among sexually active females in the total population (target range: 1.0-3.5%), (ii) gonorrhoea prevalence among FSW (2.0-6.5%), (iii) male-to-female ratio for gonorrhoea prevalence (0.60-1.00), and (iv) male-to-female ratio for all-cause symptomatic case rates (0.35-0.60; Text S1.3.1, Table S3). Target ranges for the first two outcomes came from a meta-regression of prevalence studies in Kenya between 2000-2024 (Text S1.3.2, Figure S2, Table S4). The third was from a systematic review and meta-regression in SSA,^5^ and the fourth was a DHS-based estimate.^19–22^ We generated 50,000 candidate parameter combinations from prior distributions (Table S2) using Latin Hypercube Sampling. Parameter combinations were retained if equilibrium outputs fell within all calibration ranges.

### Morbidity and health outcomes

We estimated the health burden from acute infection and longer-term sequelae with post-hoc analysis of epidemic model outcomes. In men, all symptomatic infections caused urethritis, and untreated infection could cause epididymo-orchitis.^24^ In non-pregnant women, pelvic inflammatory disease (PID) could follow any infection and progress to chronic pelvic pain (CPP), ectopic pregnancy (EP), and tubal factor infertility (TFI), individually or in combination.^24^ In pregnant women, untreated infection could cause stillbirth or result in low birth weight (LBW), neonatal pneumonia (NP), or ophthalmia neonatorum (ON),^2,3^ but not PID or its sequelae.

We used probability tree models to determine health losses following infection. We applied frameworks and assumptions from Li *et al*.^24^ for men and non-pregnant women and developed a new framework for pregnant women (Text S1.4.1, Figure S3). Health losses were measured in quality-adjusted life years (QALYs), calculated as the probability of each outcome multiplied by its duration and utility decrement. Calculations used posterior estimates of gonorrhoea incidence and duration from the epidemic model, stratified by symptom and treatment status. Acute infections (urethritis, cervicitis) were quantified as short-term decrements during the infection episode, while sequelae were quantified as longer-term losses per incident infection. PID risk increased with infection duration at an annual progression rate.^24^ Co-occurring PID sequelae (CPP, EP, TFI) were jointly modelled to capture overlapping durations and combined utility losses (Text S1.4.2).^24^

Probabilities, durations, and utility weights were from published literature and burden estimates (Table S5). We estimated stillbirth and LBW probabilities among untreated pregnant women by applying published odds ratios to background population prevalence estimates (Text S1.4.3, Table S6).^3,25,26^ Sequelae durations varied by outcome (Table S5). CPP and TFI were chronic sequelae manifesting 5 years post-infection and persisting for 10 years.^24,27^ Stillbirth-associated losses extended over life expectancy (utility weight = 0), and were thus valued equivalent to neonatal deaths.^28^ Mothers of stillborn infants or infants with adverse outcomes were assigned utility decrements for psychological impacts (Table S5).^29,30^

For infections acquired in a given year, we summed all QALY losses occurring over the infected individual’s remaining lifetime. Lifetime losses were discounted to the year of infection acquisition at 3% annually (Text S1.4.4, Table S7).

### Intervention scenarios

We modelled introduction of POCT for diagnostic confirmation or screening among five priority populations (FSW, CFSW, pregnant women, sexually active non-pregnant AGYW, and men in the total population). Strategies were modelled independently per priority population, yielding 10 scenarios, though population groups were not mutually exclusive. To assess impact under constrained availability, we assumed 25,000 POCTs annually (approximately 0.1% of adults). To assess maximum potential impact, we also modelled unrestricted POCT supply. POCTs were assumed to meet WHO optimal TPP (95% sensitivity, 98% specificity).^31^ We introduced interventions in 2025, at baseline prevalence equilibrium, and maintained them through 2030.

We compared intervention outcomes to baseline for all posterior parameter combinations. Outcomes over the 6-year intervention period were POCT coverage among the eligible population, effectiveness (proportion of infections averted or proportion of QALYs gained), and efficiency (infections averted or QALYs gained per test used). Results are reported as medians with uncertainty intervals (UIs; 2.5th–97.5th percentiles).

### Sensitivity analyses

First, to identify parameters influencing population prioritisation, we ranked priority populations by proportion of infections averted under each strategy, and calculated partial rank correlation coefficients (PRCCs) between posterior parameter values and these ranks.^32^ Second, to assess sensitivity of results to pregnancy-related sequelae assumptions, we recalculated QALY losses averted when (i) increasing the stillbirth utility weight from 0 to 0.5 and reducing LBW-associated losses from lifetime to 10 years, and (ii) increasing the stillbirth utility weight from 0 to 1 to exclude QALY losses from stillbirth. Sensitivity analyses were for constrained POCT availability scenario.

Analyses were conducted in R v4.4.1. Ordinary differential equations were implemented with *odin* (v1.5.10),^33^ Latin Hypercube Sampling with *lhs* (v1.2.0),^34^ and prevalence meta-regression for calibration outcomes with *glmmTMB* (v1.1.9).^36^

## Results

### Model calibration

Of 50,000 sampled parameter combinations, 536 were retained after calibration (Text S2.1, Figures S4-S7). At equilibrium, gonorrhoea prevalence among all sexually active 15-49-year-olds was 1.8% (UI: 1.1-3.3%; target: 1.0-3.5%) for females and 1.3% (UI:0.7-2.3%) for males. Prevalence was highest among FSW (4.7% [UI: 2.4-6.5%], target: 2.0-6.5%) and CFSW (3.6% [2.0-6.7%]), followed by pregnant women (1.9% [1.1-3.6%]) and AGYW (1.8% [1.1-3.3%]); Table S8).

### Gonorrhoea burden before intervention

We estimated 1.1 million (UI: 0.6-2.9 million) incident infections in 2025, 50.0% (45.4-55.0%) of which were among men (Figure 1A). CFSW and FSW were 7.4% (5.3-10.1%) and 1.0% (0.5-1.5%) of the total population (both sexes), respectively, and contributed 21.4% (13.7-29.3%) and 2.5% (1.0-5.6%) of total incident infections. Gonorrhoea incidence rate was highest among CFSW (13.1 [6.9-24.8] per 100 individuals) and FSW (11.9 [5.7-18.6]), and similar for pregnant women (5.0 [2.8-9.1]), AGYW (4.7 [2.8-8.5]), and total population men (4.6 [2.7-8.3]; Figure 1B). A larger share of infections were untreated among females (91.7% [87.9-94.9%]) than males (63.7% [50.6-73.8%]; Figure S8).

**Figure 1:**
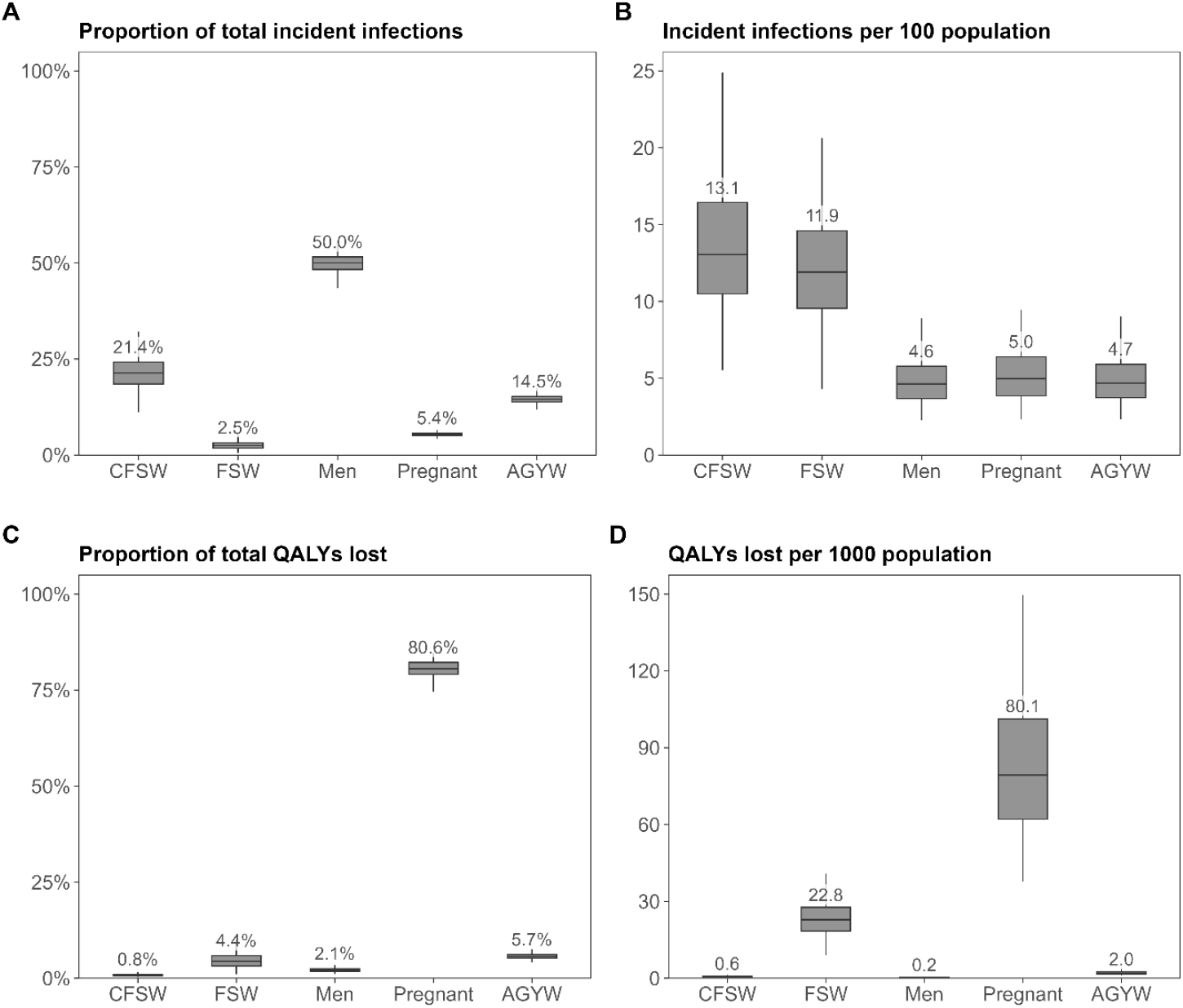
Gonorrhoea burden before intervention, 2025. Outcomes per priority population prior to intervention in 2025: (A) proportion of total incident infections, (B) incident infections per 100 individuals, (C) proportion of total lifetime QALYs lost, (D) lifetime QALYs lost per 1,000 individuals. Boxplots show median (horizontal line), interquartile range (box), and uncertainty interval (whiskers). AGYW: sexually active non-pregnant adolescent girls and young women; CFSW: clients of female sex workers; FSW: female sex workers; QALY: quality-adjusted life year.

We estimated 115,105 (66,430-207,620) total discounted lifetime QALYs lost due to infections acquired in 2025, of which 80.6% (76.1-83.8%) were among pregnant women and their infants (Figure 1C), mainly from stillbirth (70.0%) and LBW among infants (26.5%; Figure S9). QALYs lost per 1,000 individuals were highest among pregnant women (80.1 [UI: 45.1-145.0]), followed by FSW (22.8 [11.1-36.0]), AGYW (2.0 [1.2-3.7]), CFSW (0.6 [0.3-1.2]), and men (0.2 [0.1-0.5]; Figure 1C).

### Impact under constrained POCT availability

With 25,000 POCTs available annually between 2025-2030, the proportion of individuals with discharge symptoms who received diagnostic confirmation testing ranged from 8.0% (UI: 3.9-16.2%) among symptomatic total population men to 66.8% (26.0-100.0%) among FSW, and the proportion of routine healthcare attendees who received screening ranged from 0.14% (0.14-0.15%) among men to 5.8% (3.9-11.1%) among FSW (Table S9).

Diagnostic confirmation averted 2.2% (UI: 0.8-5.3%) of all incident infections between 2025-2030 when conducted among CFSW, 2.1% (0.6-5.6%) when among FSW, 1.3% (0.5-3.0%) when among men, 1.0% (0.5-2.1%) when among pregnant women, and 0.9% (0.4-2.0%) when among AGYW (Figure 2A, Table S9). The impact of screening was smaller, averting 0.6% (0.2-1.5%) of all incident infections when conducted among FSW and 0.6% (0.4-0.8%) among CFSW, followed by 0.13% (0.11-0.16%) among AGYW, 0.12% (0.10-0.13%) among men, and 0.08% (0.06-0.10%) among pregnant women (Figure 2A).

**Figure 2:**
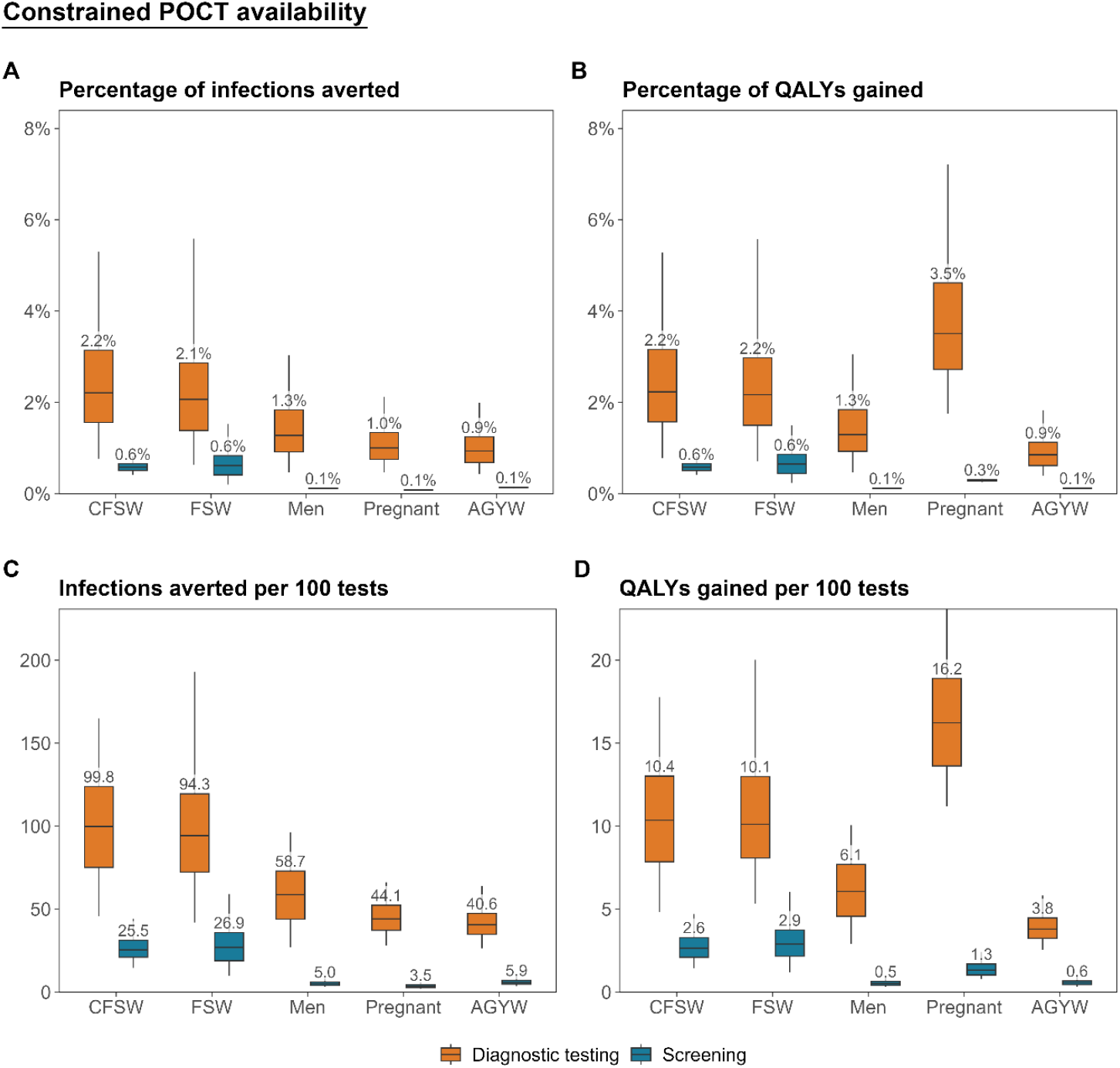
Population-level impact of diagnostic confirmation and screening strategies among each priority population under constrained POCT availability, 2025-2030. Population-level impact of deploying a constrained supply of gonorrhoea POCTs among each of five priority populations between 2025 and 2030 through either diagnostic confirmation testing of individuals accessing care for genital discharge, or screening of attendees at routine healthcare services. Impact measured as (A) percentage of incident infections averted, (B) percentage of QALYs gained, (C) incident infections averted per 100 POCTs used and (D) QALYs gained per 100 POCTs used, relative to the baseline with syndromic management only. Boxplots represent the median (horizontal line), interquartile range (box), and uncertainty interval (whiskers). AGYW: sexually active non-pregnant adolescent girls and young women; CFSW: clients of female sex workers; FSW: female sex workers; POCT: point-of-care test; QALY: quality-adjusted life year.

In contrast, the most QALYs were gained by diagnostic confirmation among pregnant women (Figure 2B, Table S9), averting 3.5% (1.8-7.2%) of population-wide lifetime QALY losses. This exceeded the impact of testing FSW (2.2% [0.7-5.6%]) or CFSW (2.2% [0.8-5.3%]), despite those strategies averting twice as many incident infections. Screening pregnant women averted 0.29% (0.25-0.34%) of QALY losses, intermediate between screening high- and lower-risk groups.

Patterns were consistent whether assessed by total impact (Figure 2A,B), or infections and QALY losses averted per test (Figure 2C,D).

### Impact under unrestricted POCT availability

With unrestricted POCTs between 2025-2030, diagnostic confirmation averted 15.3% (UI: 6.8-28.9%) of all incident infections when conducted among total population men, followed by 11.6% (4.9-24.1%) among CFSW, 6.9% (3.8-10.7%) among AGYW, 3.1% (0.7-10.0%) among FSW, and 3.0% (1.6-5.0%) among pregnant women (Figure 3A, Table S10). The relative pattern of QALYs gained across populations was similar, except diagnostic confirmation among pregnant women averted 10.7% (6.4-15.8%) of total lifetime QALY losses (Figure 3B).

**Figure 3:**
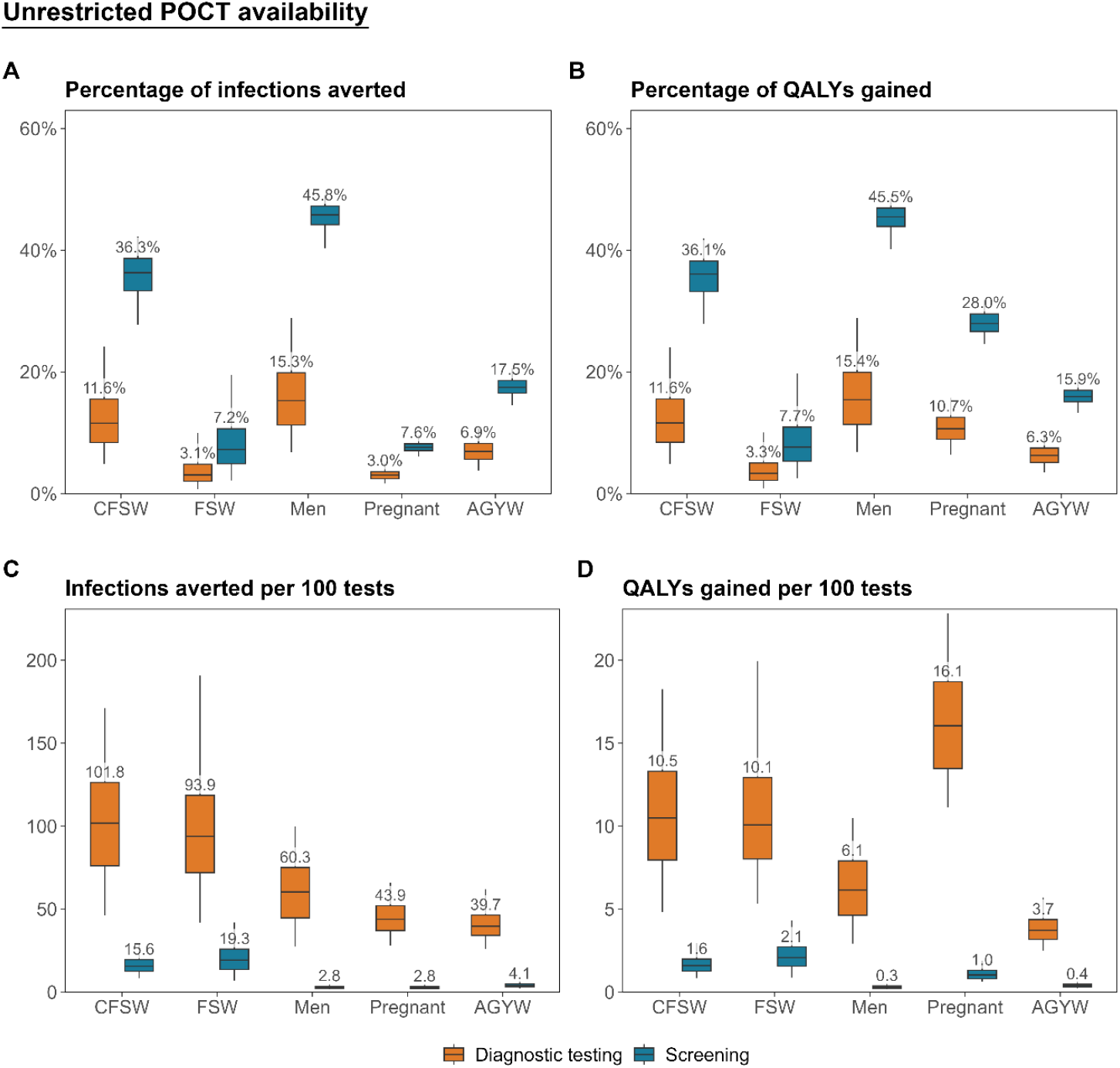
Population-level impact of diagnostic confirmation and screening strategies among each priority population under unrestricted POCT availability, 2025-2030. Population-level impact of deploying an unlimited supply of gonorrhoea POCTs among each of five priority populations between 2025 and 2030 through either diagnostic confirmation testing of individuals accessing care for genital discharge, or screening of attendees at routine healthcare services. Impact measured as (A) percentage of incident infections averted, (B) percentage of QALYs gained, (C) incident infections averted per 100 POCTs used and (D) QALYs gained per 100 POCTs used, relative to the baseline with syndromic management only. Boxplots represent the median (horizontal line), interquartile range (box), and uncertainty interval (whiskers). AGYW: sexually active non-pregnant adolescent girls and young women; CFSW: clients of female sex workers; FSW: female sex workers; POCT: point-of-care test; QALY: quality-adjusted life year.

Screening averted more infections and QALY losses than diagnostic confirmation across all populations (Figure 3A,B; Table S10) but was consistently less efficient (Figure 3C,D). Screening male populations had the largest impact, averting 45.8% (40.3-49.1%) of infections and 45.5% (40.1-48.5%) of QALY losses when conducted among total population men, and 36.3% (27.7-42.2%) and 36.1% (27.9-41.9%) when among CFSW, respectively. However, efficiency was far lower for men (2.8 [1.7-4.9] infections averted and 0.3 [0.2-0.5] QALYs gained per 100 tests) than CFSW (15.6 [8.5-28.6] and 1.6 [0.9-3.0]). Screening pregnant women averted 28.0% (24.6-31.8%) of QALY losses but with lower efficiency than higher-risk groups.

Diagnostic confirmation efficiency was similar under constrained and unrestricted POCT availability across all populations, whereas screening efficiency was consistently higher under constrained availability (Figures 3 and 4).

### Sensitivity analyses

When prioritising populations for diagnostic confirmation based on total infections averted, only 24% (n=131/536) of parameter combinations ranked populations in the same order of impact as the median estimates. However, in 89% of parameter combinations either CFSW (n=299/536, 55.8%) or FSW (n=234/536, 43.7%) were most frequently identified as the highest-impact population for testing. Prioritisation was strongly associated with the sensitivity of urethral discharge syndrome management: higher sensitivity reduced the marginal benefit of POCTs for male populations (PRCC: 0.75 [UI: 0.72-0.79] total population men; 0.67 [0.62-0.72] CFSW), and correspondingly increased the relative prioritisation of female populations (Figure S10A). For screening, 53% (n=282/536) of parameter combinations yielded the same population ranking as median estimates, with either FSW (n=297/536, 55.4%) or CFSW (n=239/536, 44.6%) most frequently identified as highest impact. Higher relative numbers of sexual partnerships among FSW increased prioritisation of FSW (-0.69 [-0.75; -0.66]), while decreasing that of CFSW (0.69 [0.65-0.74]; Figure S10B).

Pregnant women remained the highest-impact population for maximising QALYs gained under alternative health loss parameter assumptions (Figure S11). Diagnostic confirmation among pregnant women averted 3.0% (1.5-6.2%) of QALY losses under conservative parameter values, and 2.6% (1.3-5.4%) of QALY losses when stillbirth losses were excluded.

## Discussion

We evaluated how a constrained supply of gonorrhoea POCTs could be allocated across testing strategies and priority populations in Kenya to maximise epidemiologic and health impact. Introducing 25,000 POCTs annually (sufficient to test 0.1% of adults) achieved the largest overall morbidity reduction when used for diagnostic confirmation testing among symptomatic pregnant women (3.5% of QALY losses averted). Diagnostic confirmation averted the most transmission when conducted among symptomatic FSW (2.1% of incident infections) or their male clients (2.2%), though with smaller morbidity reductions than focusing on pregnant women. Screening routine health service attendees was less effective (<0.6% of infections or QALY losses averted) than diagnostic confirmation for all priority populations. With unrestricted POCTs, screening yielded larger absolute effects, particularly among total population men and CFSW, but had lower per-test returns than diagnostic confirmation.

A constrained POCT supply should be prioritised for diagnostic confirmation over screening, supporting WHO guidance to strengthen aetiologic diagnosis within syndromic management. Diagnostic confirmation was more efficient than screening because symptomatic care attendees had higher per-test positivity than routine service attendees. With unrestricted tests, screening averted more total infections and QALY losses by detecting asymptomatic infections that would otherwise remain untreated, but it required substantially more tests to achieve these gains and had lower per-test returns. The benefit of unrestricted screening was greatest among larger population groups, but is unlikely to be feasible during early implementation.

Programmes aiming to maximise morbidity reduction with constrained POCTs should prioritise diagnostic confirmation among symptomatic pregnant women. Testing this group yielded the most health gains per test used, despite relatively low infection incidence, as untreated infections in pregnancy caused disproportionate QALY losses. However, focusing exclusively on pregnant women limits opportunities for broader infection control. In settings with increasing gonorrhoea prevalence, strategies targeting incidence reduction may therefore take precedence, whereas in stable or declining epidemics, prioritising high-burden groups offers greater immediate health gains.

Programmes aiming to maximise incidence reduction with constrained POCTs should prioritise diagnostic confirmation among higher-risk populations with symptomatic infection. Testing FSW or their clients was more efficient than testing lower-risk groups, due to higher prevalence and greater indirect benefits from reduced onward transmission. Simultaneously targeting FSW and CFSW may however yield diminishing returns at high coverage due to overlapping sexual networks. As POCT availability increases and coverage among high-risk groups saturates, our results suggest that diagnostic confirmation should expand to lower-risk symptomatic males. We evaluated interventions delivered alternately to each population; future modelling should assess combined and sequential test allocation.

Diagnostic confirmation averted more infections among male than female subgroups, despite the already high sensitivity of syndromic management for urethral discharge (80–90%)^37^ compared to POCTs meeting the WHO Optimal TPP (95%). This reflects higher symptom presentation and care-seeking among males, together with fewer female syndromic cases attributable to gonorrhoea. Multiplex POCTs may have higher diagnostic utility for females, since vaginal discharge often has multiple aetiologies. As new platforms approach market introduction, their performance and optimal targeting should be re-evaluated.

Screening asymptomatic populations offered limited benefit, especially in low-prevalence groups. Even highly sensitive and specific POCTs yield low positive predictive values in such contexts, leading to false positives and unnecessary treatment, which may collaterally increase risk of AMR. Although we did not model false-positive treatment or resistance dynamics, evidence suggests intensified screening can drive resistance through greater antibiotic use.^38^ Conversely, diagnostic confirmation would reduce overtreatment,^14^ as syndromic management has poor specificity for both urethral (60-70%)^37^ and vaginal discharge (55–60%).^8^ These factors support prioritising POCTs for diagnostic use, especially where overtreatment and antibiotic exposure are common. WHO TPPs for POCTs detecting gonorrhoea resistance markers could enable more targeted treatment.^31^ Future models should incorporate resistance dynamics to align POCT deployment with antimicrobial stewardship.

Data limitations affected both calibration and parameterisation of the transmission model. Population-representative gonorrhoea prevalence estimates were scarce, and data for FSW largely came from urban cohort studies. Gaps in prevalence data required calibration based on male-to-female ratios for men and CFSW, and prevented direct calibration for AGYW and pregnant women. Sexual behaviour and care-seeking parameters relied on self-reported data, which are subject to reporting biases.^39^ Parameters for CFSW and intermediate risk groups relied on sparse data or assumptions.

Structural assumptions in the transmission model may have influenced intervention impact estimates. We assumed sexual mixing proportionate to numbers of sexual contacts, which may have under-represented the concentration of transmission in younger and higher-risk populations.^40^ We also did not incorporate heterogeneity in the frequency and exclusivity of mixing between FSW and CFSW. These assumptions are unlikely to have affected prioritisation rankings of FSW, CFSW, or pregnant women given large and contrasting differentials between these groups in incidence and morbidity. We excluded post-infection immunity, for which evidence is inconsistent,^41–43^ enabling rapid reinfection that may have underestimated the impact of POCTs on transmission.^43^ We assumed diagnosis depended solely on diagnostic sensitivity and was followed by universal access to effective treatment, overlooking barriers such as stock-outs or informal antibiotic use,^44^ which may have overestimated the incremental benefit of POCTs over syndromic management. Our modelled screening scenarios did not incorporate delivery constraints, such as outreach to identify CFSW or ANC screening occurring at a single visit rather than uniformly across gestation, which could affect the relative costs and feasibility of intervention delivery. We assumed stable baseline gonorrhoea prevalence and incidence pre-intervention, a simplification of longer-term trends but consistent with relatively stable estimates in Eastern Africa.^5^ Although biological and behavioural factors suggest AGYW may have higher prevalence than older women, limited and mixed evidence across SSA precluded age differentiation,^45–47^ potentially underestimating the impact of testing AGYW. MSM were not included despite high gonorrhoea prevalence in Kenya,^48^ though our findings for other high-risk populations suggest prioritising MSM would be efficient for reducing transmission but less for averting population health losses. Finally, behavioural responses to POCT intervention, such as partner notification, altered care-seeking, or stigma were not modelled, though could affect uptake and effectiveness.

Estimating gonorrhoea-related health losses required several assumptions. Sequelae probabilities were primarily derived from chlamydia studies, potentially misrepresenting gonorrhoea-specific risks.^24^ Several pregnancy-related outcomes, such as premature rupture of membranes and preterm birth, were excluded due to complex interactions with modelled sequelae and insufficient evidence for parameterisation,^2,3^ likely underestimating the total burden. For stillbirths, we assigned a utility loss equivalent to neonatal death, though quantification lacks consensus:^28^ some studies exclude stillbirth, while others assign lower losses to reflect the gradual accrual of life potential in early childhood.^28,49^ Our sensitivity analyses applying these alternatives did not however change the prioritisation of pregnant women for reducing morbidity.

In conclusion, gonorrhoea POCTs meeting the WHO Optimal TPP should be deployed for diagnostic confirmation rather than screening when supply is constrained in Kenya, supporting WHO guidance to strengthen aetiologic diagnosis within syndromic management.^6^ Programmes should prioritise symptomatic pregnant women to maximise morbidity reduction or high-incidence populations such as FSW and their clients to maximise transmission reduction, depending on programmatic objectives. POCTs were more efficient for diagnosing urethral than vaginal discharge, even though vaginal discharge syndrome has poorer performance for gonorrhoea diagnosis. As new POCTs approach market introduction, particularly those with multiplex or resistance detection capabilities, prioritisation strategies should be re-evaluated to reflect diagnostic performance, epidemic dynamics, and the potential for combined targeting approaches.

## Data Availability

All data produced in the present study are available upon reasonable request to the authors.

## Contribution

AC, JM, JWI-E, and M-CB conceptualised the study. JM developed the transmission model and probability trees with input from AC, JWI-E and M-CB. JM collated data, conducted literature reviews for model parameterisation, and performed all analyses. JK and PB provided expertise on local STI programme implementation and epidemiology, informing model development and contextualisation of results. JM wrote the first draft of the manuscript, which was revised with input from all authors. All authors approved the final manuscript.

