## Supplementary file for "Optimal deployment of gonorrhoea point-of-care tests: modelling the potential impact of diagnostic confirmation testing and screening strategies across five priority populations in Kenya"

#### Supplementary material

Julia Michalow[1*], Anne Cori[1], Joshua Kimani[2], Parinita Bhattacharjee[2,3], Prof Marie-Claude Boily[1], Jeffrey W. Imai-Eaton[1,3]

[1] MRC Centre for Global Infectious Disease Analysis, School of Public Health, Imperial College London, London, United Kingdom

[2] Partners for Health and Development in Africa (PHDA), Nairobi, Kenya

[3] Institute for Global Public Health, University of Manitoba, Winnipeg, Manitoba, Canada

[4] Center for Communicable Disease Dynamics, Department of Epidemiology, Harvard T.H. Chan School of Public Health, Boston, MA, USA

**Bibliography** **35**

S11 Influence of parameter assumptions for pregnancy-related sequelae on the predicted population-level impact of diagnostic confirmation and screening strategies, 2025–2030 34

S9 Predicted population-level impact of diagnostic confirmation testing and screening strategies among each priority population with constrained POCT availability, 2025–2030 30

S10 Predicted population-level impact of diagnostic confirmation testing and screening strategies among each priority population with unrestricted POCT availability, 2025–2030 31

### 1 Supplementary methods

#### 1.1 Transmission model details

The modelled population was stratified by sex (male and female), sexual risk (low, intermediate, and high, with high representing female sex workers [FSWs] and clients of female sex workers [CFSWs]), age group (15-24 years [younger] and 25-49 years [older]), and gestational status (non-pregnant and pregnant), denoted as $s \in \{m, f\}$, $r \in \{l, i, h\}$, $a \in \{y, o\}$, $g \in \{n, p\}$ throughout this supplement.

##### 1.1.1 Equations

For an individual of a given sex ($s$), sexual risk ($r$), age ($a$), and gestational status ($g$), the model is described by the following system of differential equations. In these equations, stratification indices are represented numerically as: $s \in \{0, 1\}$ for male and female; $r \in \{0, 1, 2\}$ for low, intermediate, and high sexual risk; $a \in \{0, 1\}$ for 15–24 and 25–49 years; and $g \in \{0, 1\}$ for non-pregnant and pregnant.

Parameters are defined in Table S2. In Equation 1.1, there are no terms relating to pregnancy ($f_{ra}$ and $\alpha$) as individuals have not yet become sexually active. In Equations 1.6 - 1.10, intervention-related parameters ($\delta_{srag}$ or $\tau^{h}_{srag}$) are set to zero in scenarios where diagnostic confirmation or screening interventions are not active. This allows all compartments and transitions to be retained in the model system while isolating intervention effects analytically. Intervention scenarios are detailed in Text 1.1.4.

In this chapter, $\mathbb{1}_A$ is used as an indicator function such that:

$$\mathbb{1}_A := \begin{cases} 1 & \text{if } A \text{ is true} \\ 0 & \text{if } A \text{ is false} \end{cases}$$

$$\frac{dU_{s,r,a,g}(t)}{dt} = ent_{s,r,a,g} - \psi_{s,r,a,g}\, U_{s,r,a,g} - \omega_a\, U_{s,r,a,g} + \mathbb{1}_{\{a=1\}}\, \omega_{a-1}\, U_{s,r,a-1,g} - \mu_{s,a}\, U_{s,r,a,g} \tag{1.1}$$

$$\begin{aligned} \frac{dS_{s,r,a,g}(t)}{dt} &= \psi_{s,r,a,g}\, U_{s,r,a,g} - \lambda_{s,r,a,g}\, S_{s,r,a,g} \\ &+ \pi(X_{s,r,a,g} + Y_{s,r,a,g} + Z_{s,r,a,g} + R^{y}_{s,r,a,g} + R^{h}_{s,r,a,g}) + \sigma(M^{y}_{s,r,a,g} + T^{y}_{s,r,a,g} + T^{h}_{s,r,a,g}) \\ &- \mathbb{1}_{\{s=1,g=0\}}\, f_{ra}\, S_{s,r,a,g} + \mathbb{1}_{\{s=1,g=1\}}\, f_{ra}\, S_{s,r,a,g-1} + \mathbb{1}_{\{s=1,g=0\}}\, \alpha\, S_{s,r,a,g+1} - \mathbb{1}_{\{s=1,g=1\}}\, \alpha\, S_{s,r,a,g} \\ &- \omega_a\, S_{s,r,a,g} + \mathbb{1}_{\{a=1\}}\, \omega_{a-1}\, S_{s,r,a-1,g} - \mu_{s,a}\, S_{s,r,a,g} \end{aligned} \tag{1.2}$$

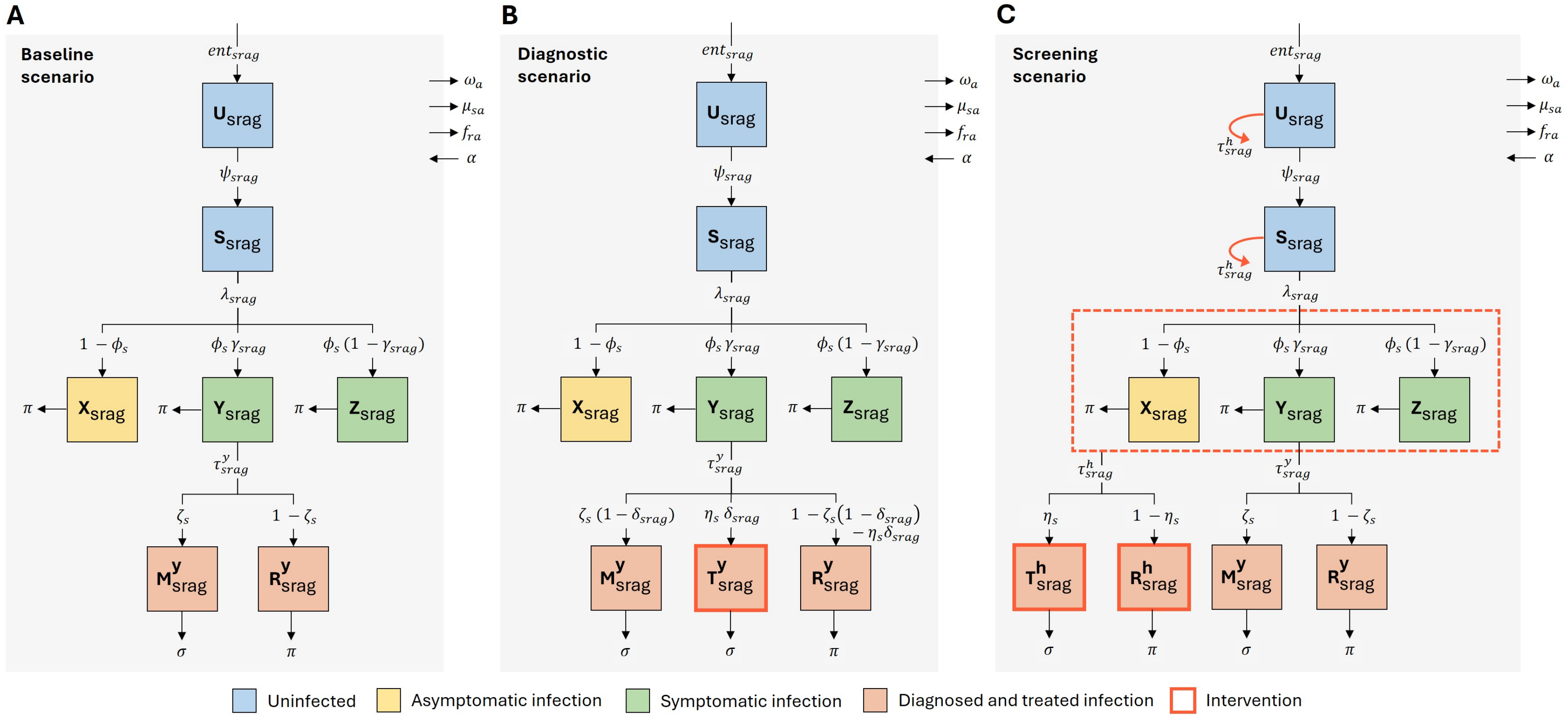

**Figure S1: Flow diagram of gonorrhoea transmission model**

Model structure showing diagnostic and treatment scenarios: (A) baseline with syndromic management, (B) diagnostic confirmation testing, and (C) screening. Population states shown as boxes: blue for uninfected ($U$: never sexually active, $S$: susceptible), yellow for asymptomatic infection ($X$), green for symptomatic infection ($Y$: with healthcare access, $Z$: without), and orange for diagnosed and treated infection ($M^y$: appropriate treatment following syndromic management, $T^y$ or $T^h$: appropriate treatment following POCT for diagnosis or screening, $R^y$ or $R^h$: inappropriate treatment). Dark orange boxes and arrows indicate POCT intervention components; the dotted box in (C) indicates that the intervention is applied to all three infection compartments. Subscripts denote stratification by sex ($s$), risk ($r$), age ($a$), and gestational status ($g$). Superscripts denote treatment among symptomatic individuals ($y$) or routine healthcare attendees ($h$). All infected individuals return to the susceptible state via natural recovery ($\pi$) or treatment ($\sigma$); these transitions are omitted for diagrammatic clarity. Outflows ($\omega_a$, $\mu_{sa}$, $f_{ra}$, $\alpha$) represent transitions or exits from relevant compartment strata due to ageing, mortality, pregnancy, or return from pregnancy. Parameters are defined in Table S2.

$$\begin{aligned}\frac{dX_{s,r,a,g}(t)}{dt} &= \lambda_{s,r,a,g}\ (1-\phi_s)\ S_{s,r,a,g} - \pi\ X_{s,r,a,g} - \tau^h_{s,r,a,g}\ X_{s,r,a,g} \\ &- \mathbb{1}_{\{s=1,g=0\}}\ f_{ra}\ X_{s,r,a,g} + \mathbb{1}_{\{s=1,g=1\}}\ f_{ra}\ X_{s,r,a,g-1} + \mathbb{1}_{\{s=1,g=0\}}\ \alpha\ X_{s,r,a,g+1} - \mathbb{1}_{\{s=1,g=1\}}\ \alpha\ X_{s,r,a,g} \\ &- \omega_a\ X_{s,r,a,g} + \mathbb{1}_{\{a=1\}}\ \omega_{a-1}\ X_{s,r,a-1,g} - \mu_{s,a}\ X_{s,r,a,g}\end{aligned} \tag{1.3}$$

$$\begin{aligned}\frac{dY_{s,r,a,g}(t)}{dt} &= \lambda_{s,r,a,g}\ \phi_s\ \gamma_{s,r,a,g}\ S_{s,r,a,g} - \pi\ Y_{s,r,a,g} - \tau^y_{s,r,a,g}\ Y_{s,r,a,g} - \tau^h_{s,r,a,g}\ Y_{s,r,a,g} \\ &- \mathbb{1}_{\{s=1,g=0\}}\ f_{ra}\ Y_{s,r,a,g} + \mathbb{1}_{\{s=1,g=1\}}\ f_{ra}\ Y_{s,r,a,g-1} + \mathbb{1}_{\{s=1,g=0\}}\ \alpha\ Y_{s,r,a,g+1} - \mathbb{1}_{\{s=1,g=1\}}\ \alpha\ Y_{s,r,a,g} \\ &- \omega_a\ Y_{s,r,a,g} + \mathbb{1}_{\{a=1\}}\ \omega_{a-1}\ Y_{s,r,a-1,g} - \mu_{s,a}\ Y_{s,r,a,g}\end{aligned} \tag{1.4}$$

$$\begin{aligned}\frac{dZ_{s,r,a,g}(t)}{dt} &= \lambda_{s,r,a,g}\ \phi_s\ (1-\gamma_{s,r,a,g})\ S_{s,r,a,g} - \pi\ Z_{s,r,a,g} - \tau^h_{s,r,a,g}\ Z_{s,r,a,g} \\ &- \mathbb{1}_{\{s=1,g=0\}}\ f_{ra}\ Z_{s,r,a,g} + \mathbb{1}_{\{s=1,g=1\}}\ f_{ra}\ Z_{s,r,a,g-1} + \mathbb{1}_{\{s=1,g=0\}}\ \alpha\ Z_{s,r,a,g+1} - \mathbb{1}_{\{s=1,g=1\}}\ \alpha\ Z_{s,r,a,g} \\ &- \omega_a\ Z_{s,r,a,g} + \mathbb{1}_{\{a=1\}}\ \omega_{a-1}\ Z_{s,r,a-1,g} - \mu_{s,a}\ Z_{s,r,a,g}\end{aligned} \tag{1.5}$$

$$\begin{aligned}\frac{dM^y_{s,r,a,g}(t)}{dt} &= \tau^y_{s,r,a,g}\ \zeta_s\ (1-\delta_{s,r,a,g})\ Y_{s,r,a,g} - \sigma\ M^y_{s,r,a,g} \\ &- \mathbb{1}_{\{s=1,g=0\}}\ f_{ra}\ M^y_{s,r,a,g} + \mathbb{1}_{\{s=1,g=1\}}\ f_{ra}\ M^y_{s,r,a,g-1} + \mathbb{1}_{\{s=1,g=0\}}\ \alpha\ M^y_{s,r,a,g+1} - \mathbb{1}_{\{s=1,g=1\}}\ \alpha\ M^y_{s,r,a,g} \\ &- \omega_a\ M^y_{s,r,a,g} + \mathbb{1}_{\{a=1\}}\ \omega_{a-1}\ M^y_{s,r,a-1,g} - \mu_{s,a}\ M^y_{s,r,a,g}\end{aligned} \tag{1.6}$$

$$\begin{aligned}\frac{dT^y_{s,r,a,g}(t)}{dt} &= \tau^y_{s,r,a,g}\ \eta_s\ \delta_{s,r,a,g}\ Y_{s,r,a,g} - \sigma\ T^y_{s,r,a,g} \\ &- \mathbb{1}_{\{s=1,g=0\}}\ f_{ra}\ T^y_{s,r,a,g} + \mathbb{1}_{\{s=1,g=1\}}\ f_{ra}\ T^y_{s,r,a,g-1} + \mathbb{1}_{\{s=1,g=0\}}\ \alpha\ T^y_{s,r,a,g+1} - \mathbb{1}_{\{s=1,g=1\}}\ \alpha\ T^y_{s,r,a,g} \\ &- \omega_a\ T^y_{s,r,a,g} + \mathbb{1}_{\{a=1\}}\ \omega_{a-1}\ T^y_{s,r,a-1,g} - \mu_{s,a}\ T^y_{s,r,a,g}\end{aligned} \tag{1.7}$$

$$\begin{aligned}\frac{dR^y_{s,r,a,g}(t)}{dt} &= \tau^y_{s,r,a,g}\ [1-\zeta_s(1-\delta_{s,r,a,g}) - \eta_s\delta_{s,r,a,g}]\ Y_{s,r,a,g} - \pi\ R^y_{s,r,a,g} \\ &- \mathbb{1}_{\{s=1,g=0\}}\ f_{ra}\ R^y_{s,r,a,g} + \mathbb{1}_{\{s=1,g=1\}}\ f_{ra}\ R^y_{s,r,a,g-1} + \mathbb{1}_{\{s=1,g=0\}}\ \alpha\ R^y_{s,r,a,g+1} - \mathbb{1}_{\{s=1,g=1\}}\ \alpha\ R^y_{s,r,a,g} \\ &- \omega_a\ R^y_{s,r,a,g} + \mathbb{1}_{\{a=1\}}\ \omega_{a-1}\ R^y_{s,r,a-1,g} - \mu_{s,a}\ R^y_{s,r,a,g}\end{aligned} \tag{1.8}$$

$$\begin{aligned}\frac{dT^h_{s,r,a,g}(t)}{dt} &= \tau^h_{s,r,a,g}\ \eta_s\ (X_{s,r,a,g} + Y_{s,r,a,g} + Z_{s,r,a,g}) - \sigma\ T^h_{s,r,a,g} \\ &- \mathbb{1}_{\{s=1,g=0\}}\ f_{ra}\ T^h_{s,r,a,g} + \mathbb{1}_{\{s=1,g=1\}}\ f_{ra}\ T^h_{s,r,a,g-1} + \mathbb{1}_{\{s=1,g=0\}}\ \alpha\ T^h_{s,r,a,g+1} - \mathbb{1}_{\{s=1,g=1\}}\ \alpha\ T^h_{s,r,a,g} \\ &- \omega_a\ T^h_{s,r,a,g} + \mathbb{1}_{\{a=1\}}\ \omega_{a-1}\ T^h_{s,r,a-1,g} - \mu_{s,a}\ T^h_{s,r,a,g}\end{aligned} \tag{1.9}$$

$$
\begin{aligned}
\frac{dR^h_{s,r,a,g}(t)}{dt} &= \tau^h_{s,r,a,g}\ (1-\eta_s)\ (X_{s,r,a,g}+Y_{s,r,a,g}+Z_{s,r,a,g}) - \pi\ R^h_{s,r,a,g} \\
&- \mathbb{1}_{\{s=1,g=0\}}\ f_{ra}\ R^h_{s,r,a,g} + \mathbb{1}_{\{s=1,g=1\}}\ f_{ra}\ R^h_{s,r,a,g-1} + \mathbb{1}_{\{s=1,g=0\}}\ \alpha\ R^h_{s,r,a,g+1} - \mathbb{1}_{\{s=1,g=1\}}\ \alpha\ R^h_{s,r,a,g} \\
&- \omega_a\ R^h_{s,r,a,g} + \mathbb{1}_{\{a=1\}}\ \omega_{a-1}\ R^h_{s,r,a-1,g} - \mu_{s,a}\ R^h_{s,r,a,g}
\end{aligned} \tag{1.10}
$$

The total ($N$), sexually active ($\hat{N}$), and infected ($I$) population are calculated as:

$$N_{s,r,a,g} = U_{s,r,a,g} + S_{s,r,a,g} + I_{s,r,a,g} \tag{1.11}$$

$$\hat{N}_{s,r,a,g} = S_{s,r,a,g} + I_{s,r,a,g} \tag{1.12}$$

$$I_{s,r,a,g} = X_{s,r,a,g} + Y_{s,r,a,g} + Z_{s,r,a,g} + M^y_{s,r,a,g} + T^y_{s,r,a,g} + R^y_{s,r,a,g} + T^h_{s,r,a,g} + R^h_{s,r,a,g} \tag{1.13}$$

##### 1.1.2 Sexual mixing

Sexual mixing was implemented as fully proportionate. The mixing matrix ($\rho_{sragsʼr'a'g'}$) represented the probability that an individual of type $s, r, a, g$ selects a sexual partner of type $s', r', a', g'$, conditional on forming a new partnership. This probability was proportional to the total number of partnerships offered by the opposite sex, as determined by their sexually active population size ($\hat{N}_{s'r'a'g'}$) and their average annual rate of partner acquisition ($c_{s'r'a'g'}$). The mixing matrix also incorporated structural constraints in the sexual network ($\theta_{srags'r'a'g'}$): (1) partnerships were only possible between individuals of the opposite sex, and (2) FSW formed partnerships exclusively with their clients (high-risk males), while clients could form partnerships with females from all risk groups.

$$\rho_{srags'r'a'g'} = \frac{\theta_{srags'r'a'g'}\ c_{s'r'a'g'}\ \hat{N}_{s'r'a'g'}}{\sum_{r''=l}^{h}\sum_{a''=y}^{o}\sum_{g''=n}^{p}\theta_{srags''r''a''g''}\ c_{s''r''a''g''}\ \hat{N}_{s''r''a''g''}} \tag{1.14}$$

$$
\theta_{srags'r'a'g'} = \begin{cases}
0 & \text{Same-sex partnerships} \\
 & \quad s = s' \\
0 & \text{FSW and non-client partnerships} \\
 & \quad s=f,\ r=h,\ s'=m,\ r'\in\{l,i\} \\
 & \quad s=m,\ r\in\{l,i\},\ s'=f,\ r'=h \\
1 & \text{Other partnerships}
\end{cases} \tag{1.15}
$$

To ensure consistency in the number of partnerships formed between sexes, a balancing procedure was applied to the annual rates of partner acquisition.[1] This adjustment was necessary because reported sexual partner numbers often differ between men and women. The balancing factor ($B_{srags'r'a'g'}$)

quantified the degree of imbalance in the total number of partnerships formed by individuals of type $s, r, a, g$ with those of type $s'r'a'g$:

$$B_{srags'r'a'g'} = \frac{c_{srag}\ \rho_{srags'r'a'g'}\ \hat{N}_{srag}}{c_{s'r'a'g'}\ \rho_{s'r'a'g'srag}\ \hat{N}_{s'r'a'g'}} \tag{1.16}$$

A value of $B_{srags'r'a'g'} = 1$ indicated perfect balance, while values greater or less than 1 indicated that individuals of sex $s$ are forming more or fewer partnerships with individuals of sex $s'$, respectively. To preserve relative differences in partner acquisition across groups while ensuring symmetry in the total number of partnerships formed between any two strata, the initial rates were adjusted as follows:

$$c^*_{srags'r'a'g'} = c_{srag}\ B^{-0.5}_{srags'r'a'g'} \tag{1.17}$$

$$c^*_{s'r'a'g'srag} = c_{s'r'a'g'}\ B^{0.5}_{srags'r'a'g'} \tag{1.18}$$

The adjusted rate ($c^*_{srags'r'a'g'}$) represented the effective annual rate at which individuals of type $s, r, a, g$ acquired new partners of type $s', r', a', g'$, after accounting for imbalances in partnership numbers between sexes. The product $c^*_{srags'r'a'g'}\rho_{srags'r'a'g'}$ therefore reflected the annual number of new partners of type $s', r', a', g'$ acquired by individuals of type $s, r, a, g$.

##### 1.1.3 Force of infection

The force of infection ($\lambda_{srag}$) determined the rate per year at which a susceptible individual of type $s, r, a, g$ acquired infection through the formation of new sexual partnerships. This depended on the per-partnership probability of transmission ($\kappa^p_{srags'r'a'g'}$), the adjusted rate of acquiring new partners ($c^*_{srags'r'a'g'}$), the probability of selecting such a partner conditional on partnership formation ($\rho_{srags'r'a'g'}$), and the prevalence of infection among potential partners ($I_{s'r'a'g'}/\hat{N}_{s'r'a'g'}$):

$$\lambda_{srag} = \sum_{r'=l}^{h} \sum_{a'=y}^{o} \sum_{g'=n}^{p} \kappa^p_{srags'r'a'g'}\ c^*_{srags'r'a'g'}\ \rho_{srags'r'a'g'}\ \frac{I_{s'r'a'g'}}{\hat{N}_{s'r'a'g'}} \tag{1.19}$$

The per-partnership transmission probability ($\kappa^p_{srags'r'a'g'}$) was determined based on the transmission probability per sex act ($\kappa^a_s$), the number of sex acts per partnership ($n^p_{srags'r'a'g'}$), the proportion of sex acts within the partnership where a condom is used ($\chi^p_{srags'r'a'g'}$), and the efficacy of condoms in preventing transmission ($e$). The formulation considered the probability of avoiding infection across all sex acts within the partnership, accounting separately for acts with and without condom use:

$$\kappa^p_{srags'r'a'g'} = 1\ -\ (1-(1-e)\kappa^a_s)^{n^p_{srags'r'a'g'}\chi^p_{srags'r'a'g'}}\ (1-\kappa^a_s)^{n^p_{srags'r'a'g'}(1-\chi^p_{srags'r'a'g'})} \tag{1.20}$$

Since data were lacking to quantify the number of sex acts within each partnership type, it was assumed that the annual number of sex acts per individual ($n$) was the same across all strata, and that sex acts were distributed equally across partners. FSW and their clients were assumed to have only one sex act per partnership. The number of sex acts per partnership was therefore defined as:

$$n^p_{srags'r'a'g'} = \begin{cases} 1 & \text{FSW and client partnerships} \\ \frac{n}{c^*_{srags'r'a'g'}\rho_{srags'r'a'g'}} & \text{Other partnerships} \end{cases} \tag{1.21}$$

To ensure consistency in condom use assumptions across partner types, the proportion of sex acts within each partnership that used a condom ($\chi^p_{srags'r'a'g'}$) was calculated by equally weighting each partner's reported condom use at last sex:

$$\chi^p_{srags'r'a'g'} = (\chi_{srag} + \chi_{s'r'a'g'})^{0.5} \tag{1.22}$$

##### 1.1.4 Intervention scenarios

Point-of-care testing for gonorrhoea was implemented from 2025 to 2030. Tests were allocated to one of five priority population groups: pregnant women (all ages and risk groups); sexually active adolescent girls and young women (AGYW; non-pregnant 15-24-year-old females who had ever had sex); FSW (all ages and gestational status); the total male population (all ages and risk groups); and CFSW (high-risk males of all ages). For each priority population, tests were used as either a diagnostic confirmation or screening intervention.

**Diagnostic confirmation:** In the diagnostic confirmation scenario, POCTs were implemented among individuals accessing care for urethral discharge and vaginal discharge syndromes. The eligible population was determined as the annual number of symptomatic cases caused by gonorrhoea ($\text{NG-case}_{srag}$) and other reproductive tract infections ($\text{RTI-case}_{srag}$), such that the same individual could be tested multiple times if they experienced repeat infections.

The number of symptomatic gonorrhoea cases accessing treatment per year ($y$) was defined as the total number of individuals leaving the $Y$ compartment, where $Y_{srag}$ represented symptomatic gonorrhoea infections in individuals planning to access treatment. This was calculated as:

$$\text{NG-case}_{srag}(y) = \int_{t=y}^{y+1} \tau^y_{srag}\, Y_{srag}(t)\, dt \tag{1.23}$$

Annual symptomatic cases due to other RTIs were calculated using the gonorrhoea aetiologic proportion

($p_s$), defined as the proportion of individuals with a given symptom who have gonorrhoea as the underlying cause. This allowed estimation of total symptomatic cases attributable to causes other than gonorrhoea by scaling observed symptomatic gonorrhoea cases accordingly. These RTI case numbers were then fixed at pre-intervention levels to ensure they remained independent of changes in gonorrhoea incidence during intervention simulations:

$$\text{RTI-case}_{srag}(y) = \begin{cases} \text{NG-case}_{srag}(y)\ \frac{(1-p_s)}{p_s} & y < 2025 \\ \text{NG-case}_{srag}(y)\ \frac{(1-p_s)}{p_s}\Big|_{y=2024} & y \geq 2025 \end{cases} \tag{1.24}$$

The annual probability of receiving a diagnostic confirmation test within a selected priority population ($\delta_T$) was calculated as the ratio of available tests to the total number of eligible symptomatic cases in that population. Each priority population $T$ comprised a subset of strata defined by sex, risk, age, and gestational status, such that $(srag) \in T$ denoted the included combinations. A maximum probability of 1 was imposed to ensure that testing coverage did not exceed the number of eligible individuals:

$$\delta_T = \min\left(1, \frac{\text{annual tests available for } T}{\sum_{(srag)\in T} \text{NG-case}_{srag}(y) + \text{RTI-case}_{srag}(y)}\right) \tag{1.25}$$

**Screening:** In the screening scenario, POCTs were implemented among individuals attending either primary healthcare or antenatal care services. The eligible population was defined as the number of individuals accessing these health services annually. Since individuals may attend care more than once per year, the annual probability of healthcare attendance ($\text{hc}_{srag}$) was converted to a rate, assuming visits follow a Poisson process with exponential spacing over time. The effective number of individuals attending healthcare services annually and eligible for screening was calculated as:

$$N^{\text{h}}_{srag} = -\log(1 - \text{hc}_{srag})(U_{srag} + S_{srag} + X_{srag} + Y_{srag} + Z_{srag}) \tag{1.26}$$

For each priority population $T$, the annual screening rate ($\tau^{\text{h}}_T$) was calculated as the ratio of available tests to the total number of eligible individuals in the priority population:

$$\tau^{\text{h}}_T = \frac{\text{annual tests available for } T}{\sum_{(srag)\in T} N^{\text{h}}_{srag}} \tag{1.27}$$

The same annual screening rate was applied to each stratum $srag \in T$.

#### 1.2 Transmission model parametrisation

##### 1.2.1 Demographic parameters

Population size was derived using age- and sex-stratified demographic estimates for the 15-49-year-old Kenyan population between 1970 and 2030 from UN World Population Prospects (WPP) 2024.[2] The female population was further stratified by gestational status. The number of pregnant women in year $y$ was estimated according to the number of births by maternal age group in the following year ($B_{fa}$) and the annual rate of delivery ($\alpha$), corresponding to a mean gestational period of 9 months (i.e. $1/\alpha = 0.75$ year):

$$N_{fap}(y) = \alpha B_{fa}(y+1) \tag{1.28}$$

Annual pregnancy rates for females were then calculated per age group as the ratio of pregnant to non-pregnant women in that group, scaled by $\alpha$ to account for pregnancy duration:

$$f_a = \alpha \frac{N_{fap}}{N_{fan}} \tag{1.29}$$

To estimate annual pregnancy rates among sexually active females, population-level rates were adjusted by the proportion sexually active in each non-pregnant female stratum ($q_{fran}$), which varied by risk group, such that:

$$f_{ra}^{model} = \frac{f_a}{q_{fran}} \tag{1.30}$$

This adjustment ensured that the total number of pregnancies was preserved. If all non-pregnant females in a stratum were sexually active ($q_{fran} = 1$), no adjustment was needed. If only a subset were sexually active ($q_{fran} < 1$), the per-capita pregnancy rate among those individuals was increased proportionally.

Each sex-age-gestational stratum was divided into sexual risk groups according to the sex-specific probability of belonging to a risk group ($\upsilon_{sr}$):

$$N_{srag}(y) = \upsilon_{sr} N_{sag}(y) \tag{1.31}$$

New male and female entrants to the population were aged 15-24 years and allocated to the $U$ compartment. The annual number of entrants was calculated using sex-stratified births 15 years prior ($B_s$) and deaths in corresponding sex and age cohorts ($D_s^{\text{age}}$) at the midpoint of exposure.[2] All entrants were non-pregnant and were stratified by risk group using the same method as for the total population.

$$ent_{srag}(y) = \mathbb{1}_{\{a=15-24,g=n\}}\, \upsilon_{sr} \left[B_s(y-15) - D_s^{0-4}(y-13) - D_s^{5-9}(y-8) - D_s^{10-14}(y-3)\right] \qquad (1.32)$$

Values were calculated for 1970 and 2030, with intermediate years interpolated linearly.

Individuals aged from 15–24 to 25–49 years and exited the 25-49 year-old group at the annual rate $\omega_a$. Sex and risk group were assumed to remain fixed over time, while gestational status changed dynamically for females based on the age- and risk-specific pregnancy rates described above.

Mortality rates ($\mu_{sa}$) from all causes were calculated by dividing total deaths by population size per sex and age in 1970 and 2030, with values interpolated linearly for intermediate years.[2]

**Table S1: Demographic parameter values**

| Stratum | Population | | Entrants | | Pregnancy rate | | Mortality rate | |
|---|---|---|---|---|---|---|---|---|
| | 1970 | 2030 | 1970 | 2030 | 1970 | 2030 | 1970 | 2030 |
| m.l.y.n | 730 072 | 4 089 571 | 96 749 | 485 610 | 0 | 0 | 0.0042 | 0.0020 |
| m.l.y.p | 0 | 0 | 0 | 0 | 0 | 0 | 0 | 0 |
| m.l.o.n | 729 196 | 6 017 108 | 0 | 0 | 0 | 0 | 0.0073 | 0.0060 |
| m.l.o.p | 0 | 0 | 0 | 0 | 0 | 0 | 0 | 0 |
| m.i.y.n | 157 253 | 880 867 | 20 839 | 104 597 | 0 | 0 | 0.0042 | 0.0020 |
| m.i.y.p | 0 | 0 | 0 | 0 | 0 | 0 | 0 | 0 |
| m.i.o.n | 157 064 | 1 296 046 | 0 | 0 | 0 | 0 | 0.0073 | 0.0060 |
| m.i.o.p | 0 | 0 | 0 | 0 | 0 | 0 | 0 | 0 |
| m.h.y.n | 154 455 | 865 197 | 20 468 | 102 736 | 0 | 0 | 0.0042 | 0.0020 |
| m.h.y.p | 0 | 0 | 0 | 0 | 0 | 0 | 0 | 0 |
| m.h.o.n | 154 270 | 1 272 990 | 0 | 0 | 0 | 0 | 0.0073 | 0.0060 |
| m.h.o.p | 0 | 0 | 0 | 0 | 0 | 0 | 0 | 0 |
| f.l.y.n | 607 808 | 3 870 978 | 101 486 | 497 481 | 0.49 | 0.19 | 0.0036 | 0.0015 |
| f.l.y.p | 132 960 | 339 634 | 0 | 0 | 0 | 0 | 0.0036 | 0.0015 |
| f.l.o.n | 735 096 | 5 828 462 | 0 | 0 | 0.32 | 0.11 | 0.0062 | 0.0043 |
| f.l.o.p | 177 531 | 487 227 | 0 | 0 | 0 | 0 | 0.0062 | 0.0043 |
| f.i.y.n | 208 485 | 1 332 056 | 34 811 | 170 642 | 0.45 | 0.17 | 0.0036 | 0.0015 |
| f.i.y.p | 45 607 | 112 233 | 0 | 0 | 0 | 0 | 0.0036 | 0.0015 |
| f.i.o.n | 252 147 | 1 999 513 | 0 | 0 | 0.32 | 0.11 | 0.0062 | 0.0043 |
| f.i.o.p | 60 895 | 166 843 | 0 | 0 | 0 | 0 | 0.0062 | 0.0043 |
| f.h.y.n | 16 827 | 107 853 | 2 810 | 13 773 | 0.41 | 0.16 | 0.0036 | 0.0015 |
| f.h.y.p | 3 681 | 8 717 | 0 | 0 | 0 | 0 | 0.0036 | 0.0015 |
| f.h.o.n | 20 351 | 161 406 | 0 | 0 | 0.32 | 0.11 | 0.0062 | 0.0043 |
| f.h.o.p | 4 915 | 13 443 | 0 | 0 | 0 | 0 | 0.0062 | 0.0043 |

Parameters derived using UN WPP 2024 estimates.[2] Values shown use median posterior estimates for the probability of being in a sexual risk group ($\upsilon_{sr}$) and the probability of being sexually active ($q_{srag}$).

##### 1.2.2 Sexual behaviour, diagnosis, and treatment parameters

Parameter values related to sexual behaviour and care-seeking for symptoms were derived from Kenya Demographic and Health Surveys (DHS) in 2003, 2008-2009, 2014 and 2022,[3–6] and Kenya Population-based HIV Impact Assessment (PHIA) in 2018.[7] For the parameters listed below, values were primarily estimated by pooling DHS and summarising data as survey-weighted proportions with design-based standard errors and 95% confidence intervals. The PHIA covered fewer relevant indicators; data were extracted from the published report as supplementary input. Variables included:

- Percentage of males who had ever paid for sex.[3,5]
- Percentage of adults who had ever had sex, stratified by sex and age group.[3–6]
- Number of sexual partners reported among sexually active adults in the past year, stratified by sex, age group, and gestational status.[3–6]
- Percentage of sexually active adults reporting condom use at last sex, stratified by sex, age group,[3–7] gestational status,[3–6] marital status,[3–7] and higher-risk sex.[3–6]
- Percentage of sexually active adults with genital discharge who reported accessing care in the past year, stratified by sex, age group, and pregnancy status.[3–6]
- Percentage of women who attended ANC at least once for their most recent pregnancy.[3–6]

**Table S2: Parameter definitions, priors, and posteriors for transmission model**

| Description | Symbol | Prior Distribution | Posterior, Median (UI) | Source |
|---|---|---|---|---|
| **Demography** | | | | |
| Initial population size in 1970 | $N^0_{srag}$ | Fixed | Table S1 | [2] |
| Annual entrants into population | $ent_{srag}$ | Fixed | Table S1 | [2] |
| Probability of being in sexual risk group | | | | |
| Male, low risk | $\upsilon_{ml}$ | Calculated | 0.70 (0.60-0.80) | $1 - \upsilon_{mi} - \upsilon_{mh}$ |
| Male, intermediate risk | $\upsilon_{mi}$ | U(0.10, 0.20) | 0.15 (0.10-0.20) | Assumption |
| Male, high risk (CFSW) | $\upsilon_{mh}$ | U(0.10, 0.20) | 0.15 (0.10-0.20) | [3–6] |
| Female, low risk | $\upsilon_{fl}$ | Calculated | 0.73 (0.67-0.79) | $1 - \upsilon_{fi} - \upsilon_{fh}$ |
| Female, intermediate risk | $\upsilon_{fi}$ | U(0.20, 0.30) | 0.25 (0.20-0.30) | Assumption |
| Female, high risk (FSW) | $\upsilon_{fh}$ | U(0.01, 0.03) | 0.02 (0.01-0.03) | [8] |
| Mean duration in age group (year) | | | | |
| 15-24 y | $1/\omega_y$ | Fixed | 10 | - |
| 25-49 y | $1/\omega_o$ | Fixed | 25 | - |
| Rate of becoming pregnant among sexually active females (year$^{-1}$) | $f_{ra}$ | Fixed | Table S1 | [2] |
| Mean duration of pregnancy (year) | $1/\alpha$ | Fixed | 0.75 | - |
| Rate of mortality (year$^{-1}$) | $\mu_{sa}$ | Fixed | Table S1 | [2] |
| **Sexual behaviour** | | | | |
| Probability of being sexually active | | | | |
| All sex, low risk, 15-24 y, non-pregnant | $q_{slyn}$ | U(0.50, 0.70) | 0.59 (0.50-0.70) | [3–6] |
| All sex, all risk, 25-49 y, non-pregnant | $q_{sron}$ | Fixed | 1 | [3–6] |
| Female, all risk, all age, pregnant | $q_{frap}$ | Fixed | 1 | - |
| Relative probability of being sexually active among all sex, 15-24 y, non-pregnant | | | | |
| Intermediate:low risk | $rq_{i:l}$ | U(1.05, 1.15) | 1.10 (1.05-1.15) | Assumption |
| High:intermediate risk | $rq_{h:i}$ | U(1.05, 1.15) | 1.10 (1.05-1.15) | Assumption |
| Rate of becoming sexually active (year$^{-1}$) | $\psi_{srag}$ | Calculated | - | -$log(1 - q_{srag})$ |
| Number of partnerships per person (year$^{-1}$) | | | | |
| All sex, low risk, 15-24 y, non-pregnant | $c_{slyn}$ | U(1.00, 1.50) | 1.24 (1.01-1.48) | [3–6] |
| Relative number of partnerships per person | | | | |
| Male, intermediate:low risk | $rc_{mi:ml}$ | U(3.00, 6.00) | 4.20 (3.08-5.87) | Assumption |
| Male, high:intermediate risk | $rc_{mh:mi}$ | U(2.50, 5.00) | 2.84 (2.05-3.89) | [3,5] |
| Female, intermediate:low risk | $rc_{fi:fl}$ | U(3.00, 6.00) | 4.44 (3.12-5.93) | Assumption |
| Female, high:intermediate risk | $rc_{fh:fi}$ | U(10.0, 25.0) | 16.4 (10.4-24.5) | [9–22] |
| 25-49:15-24 y | $rc_{o:y}$ | U(0.85, 1.00) | 0.92 (0.85-0.99) | [3–6] |
| Pregnant:non-pregnant | $rc_{p:n}$ | U(0.85, 1.00) | 0.92 (0.86-1.00) | [3–6] |
| Number of sex acts per person (year$^{-1}$) | $n$ | U(52.0, 104.0) | 79.6 (54.6-102.4) | Assumption |
| Probability of condom use per sex act | | | | |
| All sex, low risk, 15-24 y, non-pregnant | $\chi_{slyn}$ | U(0.25, 0.35) | 0.31 (0.26-0.35) | [3–6] |
| Relative probability of condom use per sex act | | | | |
| Intermediate:low risk | $r\chi_{i:l}$ | U(2.30, 2.50) | 2.40 (2.31-2.50) | [3–7] |
| High:intermediate risk | $r\chi_{h:i}$ | U(1.00, 1.15) | 1.08 (1.00-1.15) | [10–14, 16, 18, 19–23] |
| 25-49:15-24 y | $r\chi_{o:y}$ | U(0.85, 1.00) | 0.94 (0.86-1.00) | [3–6] |
| Pregnant:non-pregnant, low risk | $r\chi_{lp:ln}$ | U(0.10, 0.30) | 0.20 (0.11-0.29) | [3–6] |
| Pregnant:non-pregnant, intermediate risk | $r\chi_{ip:in}$ | U(0.10, 0.30) | 0.20 (0.10-0.30) | [3–6] |
| Pregnant:non-pregnant, high risk | $r\chi_{hp:hn}$ | U(0.85, 1.00) | 0.93 (0.86-0.99) | Assumption |
| Efficacy of condoms per sex act | $e$ | U(0.90, 1.00) | 0.95 (0.90-1.00) | [24–27] |

*Continued...*

| Description | Symbol | Prior Distribution | Posterior, Median (UI) | Source |
|---|---|---|---|---|
| **Diagnosis and treatment** | | | | |
| Probability symptomatic individuals access care | | | | |
| Male, low risk, 15-24 y, non-pregnant | $\gamma_{mlyn}$ | U(0.65, 0.75) | 0.70 (0.65-0.75) | [3–6] |
| Relative probability symptomatics access care | | | | |
| Female:male | $r\gamma_{f:m}$ | U(0.70, 0.90) | 0.81 (0.70-0.90) | [3–6] |
| Intermediate:low risk | $r\gamma_{i:l}$ | U(1.00, 1.10) | 1.05 (1.00-1.10) | Assumption |
| High:intermediate risk | $r\gamma_{h:i}$ | U(1.00, 1.10) | 1.05 (1.00-1.10) | [20–22,28,29,4,5] |
| 25-49 y:15-24 y | $r\gamma_{o:y}$ | U(1.00, 1.10) | 1.05 (1.00-1.10) | [3–6] |
| Pregnant:non-pregnant | $r\gamma_{p:n}$ | U(1.00, 1.10) | 1.05 (1.00-1.10) | [3–6] |
| Rate of treatment for symptomatic males (year$^{-1}$) | $\tau^y_m$ | U(20.0, 40.0) | 29.4 (20.6-39.4) | [30–35,36–40] |
| Relative rate of treatment among symptomatics | | | | |
| Female:male | $r\tau^y_{f:m}$ | U(0.50, 1.00) | 0.76 (0.52-0.98) | [36–40] |
| Probability of POCT diagnosis among symptomatic | $\delta_{srag}$ | Fixed | Scenario specific | Text 1.1.4 |
| Probability of primary healthcare access among all risk, 15-24 y, non-pregnant | | | | |
| Male | $h_{mryn}$ | Fixed | 0.70 | [41–45] |
| Female | $h_{fryn}$ | Fixed | 0.75 | [41–45] |
| Relative probability of primary healthcare access among all sex and risk | | | | |
| 25-49 y:15-24 y | $rh_{o:y}$ | Fixed | 1.05 | [45] |
| Pregnant:non-pregnant | $rh_{p:n}$ | Fixed | 1.20 | [3–6,46,47] |
| Rate of screening with POCT (year$^{-1}$) | $\tau^h_{srag}$ | Fixed | Scenario specific | Text 1.1.4 |
| Sensitivity of syndromic management | | | | |
| Male | $\zeta_m$ | U(0.80, 0.90) | 0.85 (0.80-0.90) | [48] |
| Female | $\zeta_f$ | U(0.40, 0.50) | 0.45 (0.40-0.50) | [49,50] |
| Sensitivity of POCT | | | | |
| Male | $\eta_m$ | Fixed | 0.95 | [51] |
| Female | $\eta_f$ | Fixed | 0.95 | [51] |
| **Natural history** | | | | |
| Probability of transmission per sex act | | | | |
| Female-to-male | $\kappa_m$ | U(0.15, 0.30) | 0.23 (0.15-0.30) | [35,52–56] |
| Male-to-female | $\kappa_f$ | U(0.10, 0.40) | 0.19 (0.10-0.37) | [32,35,54–56] |
| Probability infection is symptomatic | | | | |
| Male | $\phi_m$ | U(0.45, 0.90) | 0.60 (0.46-0.85) | [34,35,54,55,57,58] |
| Female | $\phi_f$ | U(0.20, 0.45) | 0.35 (0.22-0.44) | [34,35,54,55,57–59] |
| Rate of natural recovery (year$^{-1}$) | $\pi$ | U(1.80, 2.80) | 2.31 (1.84-2.76) | [34,35,55,60–62] |
| Rate of recovery with treatment (year$^{-1}$) | $\sigma$ | U(26.0, 52.0) | 39.0 (26.6-51.5) | [63] |
| Probability syndrome is caused by gonorrhoea | | | | |
| Male (urethral discharge) | $p_m$ | U(0.65, 0.8) | 0.73 (0.65-0.80) | [64] |
| Female (vaginal discharge) | $p_f$ | U(0.10, 0.20) | 0.14 (0.10-0.20) | [64] |

Row shading indicates fixed (light blue) or estimated (white) parameters. CFSW: clients of female sex workers, FSW: female sex workers, UI: Uncertainty interval, U: Uniform distribution (min, max), y: years.

#### 1.3 Transmission model calibration

##### 1.3.1 Calibration outcomes

Calibration outcomes included: (i) gonorrhoea prevalence among sexually active females in the general population, (ii) gonorrhoea prevalence among FSW, (iii) male-to-female ratio for gonorrhoea prevalence, and (iv) male-to-female ratio for all-cause symptomatic case rates (Table S3). Due to limited studies, gonorrhoea prevalence among males was not used as a direct calibration outcome. Instead, we used ratios for male-to-female gonorrhoea prevalence and male-to-female all-cause symptomatic case rate.

**Table S3: Calibration outcomes**

| Variable | Target | Estimate (95%CI) informing target | Source for estimate |
|---|---|---|---|
| **Gonorrhoea prevalence** | | | |
| Female, overall | U(1.0%, 3.5%) | 2.2% (1.3 - 3.5%) | Meta-regression estimate in 2020 (Figure S2) |
| Female sex workers | U(2.0%, 6.5%) | 3.7% (2.1 - 6.4%) | Meta-regression estimate in 2020 (Figure S2) |
| **Male-to-female ratios** | | | |
| Prevalence | U(0.60, 1.00) | 0.81 (0.61 - 1.09) | Meta-regression estimate in 2020[65] |
| Symptomatic case rate | U(0.35, 0.60) | 0.46 (0.36 - 0.58) | Analysis of Kenya DHS[3–5] |

Symptomatic case rate: annual number of symptomatic urethral or vaginal discharge cases due to any cause, divided by the sexually active population size.

##### 1.3.2 Prevalence estimates

Gonorrhoea prevalence was estimated through systematic data collation and meta-regression. For the general population, we extracted prevalence observations (n = 24) from Kenyan studies between 2000 and 2024, with diagnostic test performance adjustment, from our previous systematic review.[65] These studies primarily involved sexually active populations. For FSW, we conducted an additional PubMed search using the same inclusion criteria, with an added search domain for sex work (i.e. “sex work”, “sex worker”, “prostitute”, “commercial sex”). After de-duplication of identified articles, prevalence observations (n = 12) were extracted and adjusted for diagnostic test performance using the same approach as in our prior analysis (Table S4).[65]

A generalised linear mixed-effects meta-regression model was then fit to the collated data. Fixed effects included year (midpoint of data collection), population group (general population, FSW), sex (female, male), age group (younger adults 12–25 years, all adults $\geq$ 12 years), and HIV status (HIV negative, non-stratified status). Interaction terms between year and sex, and year and population group, were not included due to limited observations. A study-level random intercept accounted for between-study heterogeneity. Model predictions were used to estimate prevalence for adults in the general and FSW populations, without stratifying by HIV status (Figure S2). Predictions were not stratified by age group, due to limited observations among younger adults (n = 6 general population, n = 0 FSW).

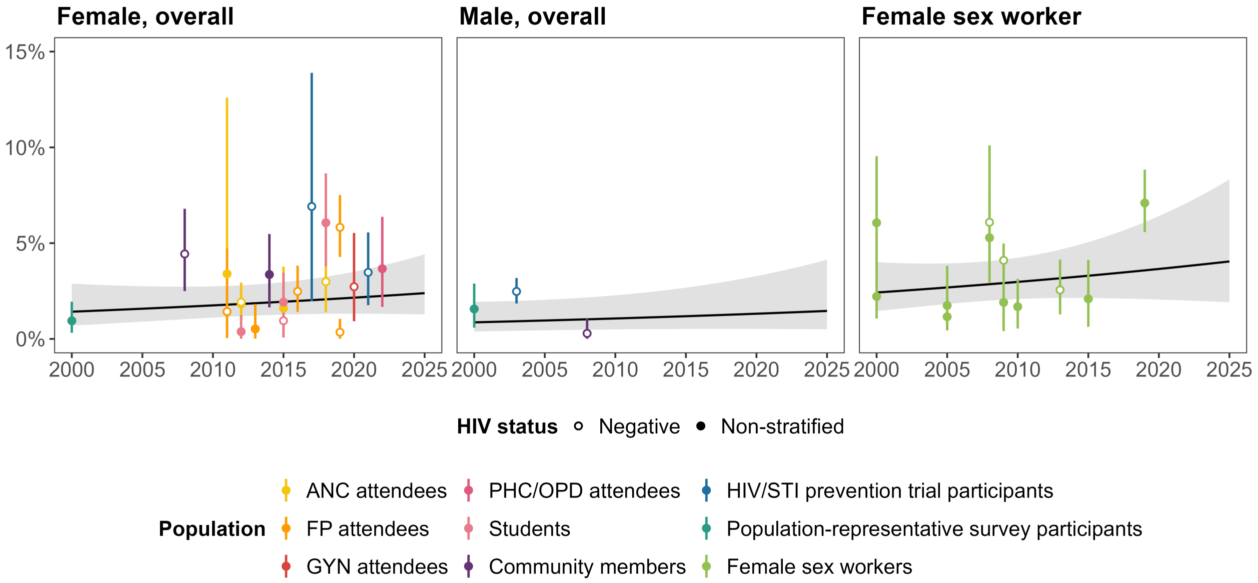

**Figure S2: Estimated prevalence of gonorrhoea among the general population and female sex workers in Kenya, 2000-2025**

Gonorrhoea prevalence among male and female general populations and FSW between 2000-2025. Lines and shading depict mean estimates with 95% CIs. Points represent study observations. ANC: antenatal care, FP: family planning, GYN: gyneacology, PHC/OPD: primary healthcare or outpatient department.

**Table S4: Study observations of gonorrhoea prevalence in Kenya**

| Sex | Population | Year | Setting | Age | HIV status | Samp | Test | Sens (%) | Spec (%) | N | Prevalence Raw (%) | Prevalence Adjusted (%, 95%CI) | Reference |
|---|---|---|---|---|---|---|---|---|---|---|---|---|---|
| **Population category: General** | | | | | | | | | | | | | |
| F | Population-representative survey participants | 2000 | Mombasa | 15 to 49 | Non-stratified | UR | NAAT | 91.6 | 100.0 | 571 | 0.7 | 0.9 (0.3-1.9) | Hawken 2002[66] |
| F | Community members | 2008 | Kisumu | 18 to 34 | Negative | GE | NAAT | 93.3 | 99.2 | 424 | 4.7 | 4.4 (2.5-6.8) | Otieno 2015[67] |
| F | ANC attendees | 2011 | Mombasa | 18 to 35 | Non-stratified | GE | NAAT | 93.3 | 99.2 | 30 | 0.0 | 3.4 (0.1-12.6) | Jespers 2014[68] |
| F | FP attendees | 2011 | Mombasa | 18 to 35 | Negative | GE | NAAT | 93.3 | 99.2 | 110 | 0.9 | 1.4 (0.0-4.7) | Jespers 2014[68] |
| F | ANC attendees | 2012 | Western Region | 14+ | Negative | GE | NAAT | 93.3 | 99.2 | 1276 | 2.5 | 1.9 (1.1-2.9) | Kinuthia 2015[69] |
| F | Students | 2012 | Western Region | 14 to 17 | Non-stratified | GE | NAAT | 93.3 | 99.2 | 511 | 0.6 | 0.4 (0.0-1.2) | Kerubo 2016[70] |
| F | FP attendees | 2013 | Nairobi | 20 to 49 | Non-stratified | GE | Culture | 75.7 | 100.0 | 249 | 0.0 | 0.5 (0.0-1.8) | Maina 2016[71] |
| F | Community members | 2014 | Kisumu | 18 to 34 | Non-stratified | GE | NAAT | 93.3 | 99.2 | 457 | 3.7 | 3.4 (1.6-5.5) | Oliver 2018[72] |
| F | ANC attendees | 2015 | Kilifi | 18 to 45 | Non-stratified | UR | NAAT | 91.6 | 100.0 | 202 | 1.0 | 1.6 (0.3-3.8) | Masha 2017[73] |
| F | Students | 2015 | Thika | 16 to 20 | Negative | GE | NAAT | 93.3 | 99.2 | 373 | 1.3 | 1.0 (0.1-2.5) | Yuh 2020[74] |
| F | FP attendees | 2016 | Kisumu | 16 to 35 | Negative | GE | NAAT | 93.3 | 99.2 | 901 | 3.0 | 2.5 (1.4-3.8) | Deese 2021[75] |
| F | Prevention trial participants | 2017 | Kisumu | 18 to 50 | Negative | GE/ UR | NAAT | 91.6 | 99.2 | 82 | 6.1 | 6.9 (2.0-13.9) | Mgodi 2021[76] |
| F | ANC attendees | 2018 | NR | NR | Negative | GE | NAAT | 93.3 | 99.2 | 474 | 3.4 | 3.0 (1.4-4.9) | Madanitsa 2023[77] |
| F | Students | 2018 | Siaya | 14 to 22 | Non-stratified | GE | NAAT | 93.3 | 99.2 | 436 | 6.2 | 6.1 (3.8-8.6) | Mehta 2023[78] |
| F | FP attendees | 2019 | Nairobi, Mombasa | 18 to 45 | Negative | GE | NAAT | 93.3 | 99.2 | 691 | 0.7 | 0.4 (0.0-1.0) | Lokken 2022[79] |
| F | FP attendees | 2019 | Kisumu | 16 to 25 | Negative | GE | NAAT | 93.3 | 99.2 | 1000 | 6.1 | 5.8 (4.3-7.5) | Celum 2022[80] |
| F | GYN attendees | 2020 | Thika, Kisumu | 15 to 30 | Negative | UR | NAAT | 91.6 | 100.0 | 198 | 2.0 | 2.7 (0.9-5.5) | Heffron 2021[81] |

*Continued...*

| Sex | Population | Year | Setting | Age | HIV status | Samp | Test | Sens (%) | Spec (%) | N | Prevalence Raw (%) | Prevalence Adjusted (%, 95%CI) | Reference |
|---|---|---|---|---|---|---|---|---|---|---|---|---|---|
| F | Prevention trial participants | 2021 | Kisumu | 18 to 30 | Negative | GE | NAAT | 93.3 | 99.2 | 448 | 3.8 | 3.5 (1.8-5.6) | Oware 2023[82] |
| F | OPD attendees | 2022 | Kitui | 15 to 44 | Non-stratified | GE | Culture | 75.7 | 100.0 | 322 | 2.5 | 3.7 (1.7-6.4) | Mbuvi 2024[83] |
| M | Population-representative survey participants | 2000 | Mombasa | 15 to 49 | Non-stratified | UR | NAAT | 80.9 | 99.9 | 583 | 1.2 | 1.6 (0.6-2.9) | Hawken 2002[66] |
| M | Prevention trial participants | 2003 | Kisumu | 18 to 24 | Negative | UR | NAAT | 80.9 | 99.9 | 2754 | 2.1 | 2.5 (1.8-3.2) | Bailey 2007[84] |
| M | Community members | 2008 | Kisumu | 18 to 34 | Negative | UR | NAAT | 80.9 | 99.9 | 422 | 0.0 | 0.3 (0.0-1.1) | Otieno 2015[67] |
| **Population category: Female sex workers** | | | | | | | | | | | | | |
| F | Female sex workers | 2000 | Nairobi | 18 to 35 | Non-stratified | GE | NAAT | 93.3 | 99.2 | 295 | 6.1 | 6.1 (3.4-9.3) | Cohen 2007[85] |
| F | Female sex workers | 2000 | Mombasa | 15 to 65 | Non-stratified | UR | NAAT | 91.6 | 100.0 | 493 | 1.8 | 2.2 (1.1-3.8) | Hawken 2002[11] |
| F | Female sex workers | 2005 | Kisumu | NR | Non-stratified | UR | NAAT | 91.6 | 100.0 | 250 | 1.2 | 1.7 (0.4-3.8) | Kwena 2010[86] |
| F | Female sex workers | 2005 | Mombasa | 16+ | Non-stratified | GE | Culture | 75.7 | 100.0 | 677 | 0.7 | 1.2 (0.5-2.3) | Chersich 2007[87] |
| F | Female sex workers | 2008 | Nairobi | 18 to 55 | Negative | GE | NAAT | 93.3 | 99.2 | 200 | 6.0 | 6.1 (2.8-10.3) | Priddy 2011[13] |
| F | Female sex workers | 2008 | Kisumu | NR | Non-stratified | GE | NAAT | 93.3 | 99.2 | 474 | 5.5 | 5.3 (3.2-7.8) | Vandenhoudt 2013[14] |
| F | Female sex workers | 2009 | Nairobi | 18 to 49 | Non-stratified | GE | NAAT | 93.3 | 99.2 | 348 | 2.3 | 1.9 (0.4-3.9) | Gomih-Alakija 2014[15] |
| F | Female sex workers | 2009 | Nairobi | NR | Negative | GE | Culture | 75.7 | 100.0 | 2931 | 3.1 | 4.1 (3.3-5.0) | Preston 2013[88] |
| F | Female sex workers | 2010 | Nairobi | 18+ | Non-stratified | GE | NAAT | 93.3 | 99.2 | 596 | 2.2 | 1.7 (0.5-3.1) | Musyoki 2015[16] |
| F | Female sex workers | 2013 | Mombasa | NR | Negative | GE | NAAT | 93.3 | 99.2 | 660 | 3.0 | 2.6 (1.3-4.1) | Willcox 2021[18] |
| F | Female sex workers | 2015 | Mombasa | 19 to 66 | Non-stratified | GE | NAAT | 93.3 | 99.2 | 399 | 2.5 | 2.1 (0.6-4.0) | Grabert 2022[89] |
| F | Female sex workers | 2019 | Nairobi | 18 to 45 | Non-stratified | UR | NAAT | 91.6 | 100.0 | 1042 | 6.4 | 7.1 (5.6-8.8) | Beksinska 2021[19] |

Study year is midpoint between start and end of data collection period. Sensitivity and specificity of diagnostic tests collated per approach outlined by Michalow *et al.* (2025).[65] ANC: antenatal care, FP: family planning, F: female, GE: genital fluid, GYN: gynaecology clinic M: male, OPD: outpatient department, samp: sample, sens: sensitivity, spec: specificity, UR: urine, NR: not reported.

#### 1.4 Quality-adjusted life years lost

##### 1.4.1 Probability trees and parametrisation

Probability tree structures for sequelae in men, non-pregnant women, and pregnant women are shown in Figure S3. Frameworks for men and non-pregnant women were from Li *et al*,[90] while a new structure was developed to capture pregnancy outcomes.

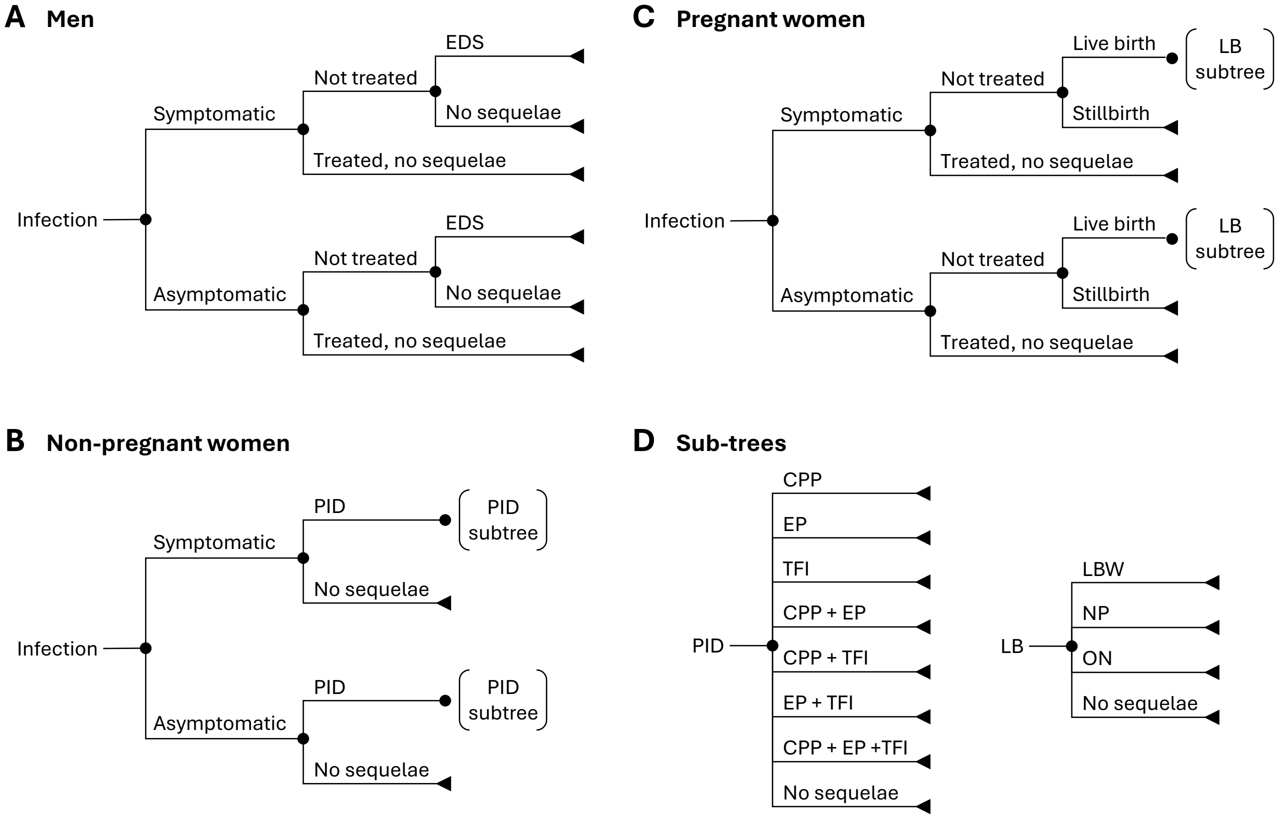

**Figure S3: Probability trees for gonorrhoea sequelae**

Probability trees for sequelae of gonorrhoea among (A) men, (B) non-pregnant women, and (C) pregnant women, with complications of PID and LB shown as separate subtrees (D). CPP: chronic pelvic pain, EDS: epididymo-orchitis, EP: ectopic pregnancy, LB: live birth, LBW: low birth weight, NP: neonatal pneumonia, OP: ophthalmia neonatorum, PID: pelvic inflammatory disease, TFI: tubal factor infertility. Trees and subtrees for men and non-pregnant women adapted from Li *et al.* 2023.[90]

Health losses were quantified as quality-adjusted life years (QALYs), where one QALY equals one year lived in perfect health. Each health state was assigned a utility value between 0 (death) and 1 (perfect health), where a utility decrement (1 - utility) represents the fraction of a healthy year lost due to that outcome. To calculate QALY losses at the population level, we used posterior estimates of gonorrhoea incidence and duration, distinguishing between episodes that were symptomatic or asymptomatic, and treated or untreated. For acute infection episodes (urethritis or symptomatic cervicitis), we multiplied person-years of infection for each subgroup by the corresponding utility decrement to capture shorter-term reductions in quality of life. For longer-term sequelae, we multiplied the number of incident infections by the probability that each sequela would result from an infection episode, the duration of the sequela, and its associated utility decrement.

Parameter values for probabilities, durations, and utilities were obtained from published literature (Table S5). Sequelae probabilities were mainly derived from longitudinal studies of chlamydia, due to sparse direct evidence for gonorrhoea.[90,91] Values for utility weights and their associated durations were from the Global Burden of Disease study,[92,93] the Institute of Medicine study,[94] and other published literature. CPP and TFI were classified as chronic sequelae with onset 5 years post-infection and duration of 10 years.[90,95] Stillbirth losses were values as equivalent to neonatal deaths (utility weight of 0 over life expectancy at birth), due to limited consensus on alternative quantification approaches.[96] Mothers with stillbirth were assigned a utility reduction for 10 years to reflect long-term psychological impacts.[97] Utility decrements were also included for mothers of infants with LBW, NP, or ON.[98]

**Table S5: Parameters for the probability, duration, and utility of gonorrhoea sequelae**

| Sequelae | Probability per infection episode | Duration (years) | Utility weight |
|---|---|---|---|
| **Men** | | | |
| Urethritis, given symptomatic infection | 1.000 [90] | Simulated* | 0.961 [92,94] |
| Urethritis, given asymptomatic infection | 0 [90] | Simulated* | 1.000 [92] |
| EDS, given untreated infection | 0.043 [90†] | 0.019 [94] | 0.665 [92,94] |
| **Non-pregnant women** | | | |
| Symptomatic infection | Simulated* | Simulated* | 0.994 [92] |
| Asymptomatic infection | Simulated* | Simulated* | 1.000 [92] |
| PID | Simulated with annual rate ‡ | 0.028 [94] | 0.756 [92,94] |
| CPP, given PID | 0.261 [90†] | 10.00 [90,95] | 0.759 [92,94] |
| TFI, given PID | 0.168 [90†] | 10.00 [90,95] | 0.905 [92,94] |
| EP, given PID | 0.070 [90†] | 0.078 [94] | 0.731 [92,94] |
| **Pregnant women** | | | |
| Symptomatic infection | Simulated* | Simulated* | 0.994 [92] |
| Asymptomatic infection | Simulated* | Simulated* | 1.000 [92] |
| Stillbirth, given untreated infection | 0.047 ‖ | Neonatal: 64.00 [2]<br>Maternal: 10.00 [97] | Neonatal: 0<br>Maternal: 0.862 [97] |
| LBW, given LB with untreated infection | 0.176 ‖ | Neonatal: 64.00 [2]<br>Maternal: 10.00 § | Neonatal: 0.894 [93]<br>Maternal: 0.999 § |
| NP, given LB with untreated infection | 0.160 [99†] | 0.167 [94] | Neonatal: 0.790 [94]<br>Maternal: 0.999 § |
| ON, given LB with untreated infection | 0.180 [99†] | 0.500 [94] | Neonatal: 0.970 [94]<br>Maternal: 0.999 § |

Utilities with two sources were averaged. *Probability and duration of symptomatic and asymptomatic gonorrhoea varied per posterior parameter combination. †Probabilities of sequelae approximated from studies of chlamydia, due to limited gonorrhoea-specific data.[90] ‡Probability of PID calculated using mean annual progression rate of 0.15 per year[100] and posterior estimates for duration of infection.[90] ‖Probabilities of stillbirth and LBW among pregnant women with untreated gonorrhoea estimated by combining odds ratios from meta-analyses with background prevalence in the general population of pregnant women.[101–103] §Assumption.[98] CPP: chronic pelvic pain, EDS: epididymitis, EP: ectopic pregnancy, LB: live birth, LBW: low birth weight, NP: neonatal pneumonia, ON: ophthalmia neonatorum, PID: pelvic inflammatory disease, TFI: tubal factor infertility.

##### 1.4.2 Equations for individual and combined sequelae

QALYs lost were calculated separately for individual and combined sequelae. In the notation used below, subscripts $s$ and $\ell$ refer to shorter- and longer-term sequelae, respectively. For individual sequelae (whether short- or long-term), QALYs lost ($Q$) were calculated as the product of the number of incident gonorrhoea cases ($x$), the probability of developing the sequela ($p$), the duration of utility loss ($d$), and the decrement in health utility $(1-u)$:

$$Q = x\,p\,d\,(1-u) \tag{1.33}$$

For combined sequelae following PID, QALYs lost were calculated in two parts, consistent with the approach used by Li *et al.*[90] covering: (i) the overlapping period, during which both sequelae occurred simultaneously, corresponding to the duration of the shorter-term sequela ($d_s$); and (ii) the extended period, during which only the longer-term sequela persisted, defined as the remaining duration of the longer-term sequela ($d_\ell - d_s$ where $d_\ell > d_s$). During the overlapping period, the joint utility was calculated as the product of the individual utility weights ($u_s\,u_\ell$). During the extended period, only the utility of the longer-term sequela ($u_\ell$) was applied. The total QALY loss among individuals with combined sequelae was:

$$Q_{s\ell} = x\ p_s\ p_\ell\ \left[(1-u_s\,u_\ell)\,d_s + (1-u_\ell)(d_\ell - d_s)\right] \tag{1.34}$$

Here, $x$ denoted the estimated number of PID cases, $p_s$ and $p_\ell$ were the probabilities of developing each sequela, $u_s$ and $u_\ell$ were the respective utility weights, and $d_s$ and $d_\ell$ were the corresponding durations.

For sequelae extending beyond the year of infection, durations were calculated using a life table approach and were discounted to the year of infection.[90]

##### 1.4.3 Adverse birth outcomes

To quantify QALYs lost due to stillbirth and LBW, estimates were required for the probability of each outcome among pregnant women with untreated gonorrhoea. However, direct estimates of these probabilities are rarely available. Most studies report associations using odds ratios (ORs), which quantify the relative odds of an outcome in exposed versus unexposed groups, i.e., among women with untreated gonorrhoea compared to those without gonorrhoea or with treated infection. ORs alone cannot estimate absolute risks, since they do not account for underlying prevalence of the outcome in the general population. Therefore, we used the approach described by Nyemba *et al.*[104] to convert ORs into absolute probabilities suitable for QALY calculations, by linking relative and absolute measures through algebraic relationships.

The approach involved expressing the outcome probability among exposed individuals ($p_1$) and unexposed individuals ($p_0$) using both the OR and risk ratio (RR):

$$\text{OR} = \frac{p_1}{1-p_1} / \frac{p_0}{1-p_0} \tag{1.35}$$

$$\text{RR} = \frac{p_1}{p_0} \tag{1.36}$$

To express RR in terms of OR, Equation 1.35 was substituted into 1.36:

$$\text{RR} = \frac{\text{OR}}{1 - p_0(1 - \text{OR})} \tag{1.37}$$

The background prevalence of the adverse outcome in the general pregnant population ($u$) was calculated using the median equilibrium prevalence of untreated gonorrhoea at baseline ($g$):

$$u = g \cdot p_1 + (1 - g) \cdot p_0 \tag{1.38}$$

Substituting Equation 1.37 and solving for $p_0$ yielded:

$$p_0 = \frac{u}{1 + g \cdot (\frac{\text{OR}}{1-p_0(1-\text{OR})} - 1)} \tag{1.39}$$

Equation 1.39 was rearranged into a quadratic Equation:

$$a \cdot p_0^2 + b \cdot p_0 + c = 0 \tag{1.40}$$

Where the coefficients were defined as:

$a = (1 - g)(\text{OR} - 1)$

$b = 1 - (\text{OR} - 1)(u - g)$

$c = -u$

The value for $p_0$ was obtained using the quadratic formula: $p_0 = \frac{-b \pm \sqrt{b^2 - 4ac}}{2a}$

The value for $p_1$ was then calculated using Equation 1.35.

The odds ratios, background prevalence, and resulting probability estimates ($p_1$) are in Table S6.

**Table S6: Probability of adverse outcomes in pregnant women with untreated gonorrhoea.**

| **Parameter** | **Stillbirth** | **Low birth weight** |
|---|---|---|
| Odds ratio (effect of gonorrhoea on outcome) | 2.16 (1.25-3.46)[103] | 1.66 (1.12 - 2.48)[103] |
| Background prevalence of outcome in general pregnant population | 0.023[101] | 0.115[102] |
| Estimated probability of outcome in pregnant women with untreated gonorrhoea | 0.047 | 0.176 |
| Estimated probability of outcome in pregnant women with treated gonorrhoea or no gonorrhoea | 0.023 | 0.114 |

##### 1.4.4 Duration and discounting

Long-term durations were assigned to chronic sequelae of PID (CPP and TFI) in non-pregnant women and adverse birth outcomes (stillbirth and LBW) in pregnant women and their infants. For chronic sequelae, a 10-year duration was assumed, representing a conservative intermediate between the 5-year, 10-year, and lifetime durations considered by Li *et al.*[90] Stillbirth and LBW losses were valued using life expectancy at birth (65 years).[2,93,105–107] For mothers, stillbirth-associated losses were valued over 10 years based on maternal perinatal mortality surveys,[97] and LBW-associated maternal impacts were assumed to have the same 10-year duration.

Years lived with long-term sequelae were estimated assuming infections and sequelae onset at age group midpoints: age 20 for the 15-24 year group and age 37 for the 25-49 years group. For chronic sequelae, onset was modelled with a 5 year lag after infection, with health losses occurring from ages 25-35 years for the younger group and 42-52 years for the older group. Years lived were calculated using a life table approach with discounting applied from the year of infection.[90] Life tables were constructed using five-year age intervals for the conditional probabilities of all-cause mortality and survival from UN WPP projections.[2] Deaths were assumed to occur at the midpoint of each age interval. Age weights were not applied, per GBD and WHO conventions valuing health losses equally across age.[108,109]

For each interval, the undiscounted years lived ($\mathrm{YL}_u$) were calculated by incorporating the probability of surviving from age at infection to the start of the age interval ($p_i$), and the probability of death during the interval ($p_d$).

$$\mathrm{YL}_u = p_i\,[2.5\,p_d + 5\,(1 - p_d)] \tag{1.41}$$

The years lived were then discounted ($\mathrm{YL}_d$) for each interval using a 3% annual discount rate applied from infection onset to the interval midpoint:

$$\mathrm{YL}_d = \sum_{k=0}^{n-1} \frac{1}{(1+r)^{t+k}} \tag{1.42}$$

Where $r$ was the annual discount rate, $t$ was the number of years from infection onset to interval start (including sequelae onset delays), $n$ was the number of years with the sequela, and $k$ indexed each year within the interval.

Undiscounted and discounted years lived were summed across relevant age intervals (Table S7) and used to calculate lifetime QALYs lost due to each sequela.

**Table S7: Years lived with health losses due to each sequela**

| Sequelae | Age group | Age of infection | Duration sequela | Years lived | |
|---|---|---|---|---|---|
| | | | | Undiscounted | Discounted |
| Chronic: CPP and TFI | 15-24 years | 20 | 10 | 9.75 | 7.28 |
| Chronic: CPP and TFI | 25-49 years | 37 | 10 | 9.23 | 6.92 |
| Maternal: Stillbirth and LBW | 15-24 years | 20 | 10 | 9.89 | 8.56 |
| Maternal: Stillbirth and LBW | 25-49 years | 37 | 10 | 9.59 | 8.46 |
| Stillbirth and LBW | Neonate | 0 | 64 | 56.5 | 26.3 |

### 2 Supplementary results

#### 2.1 Transmission model calibration

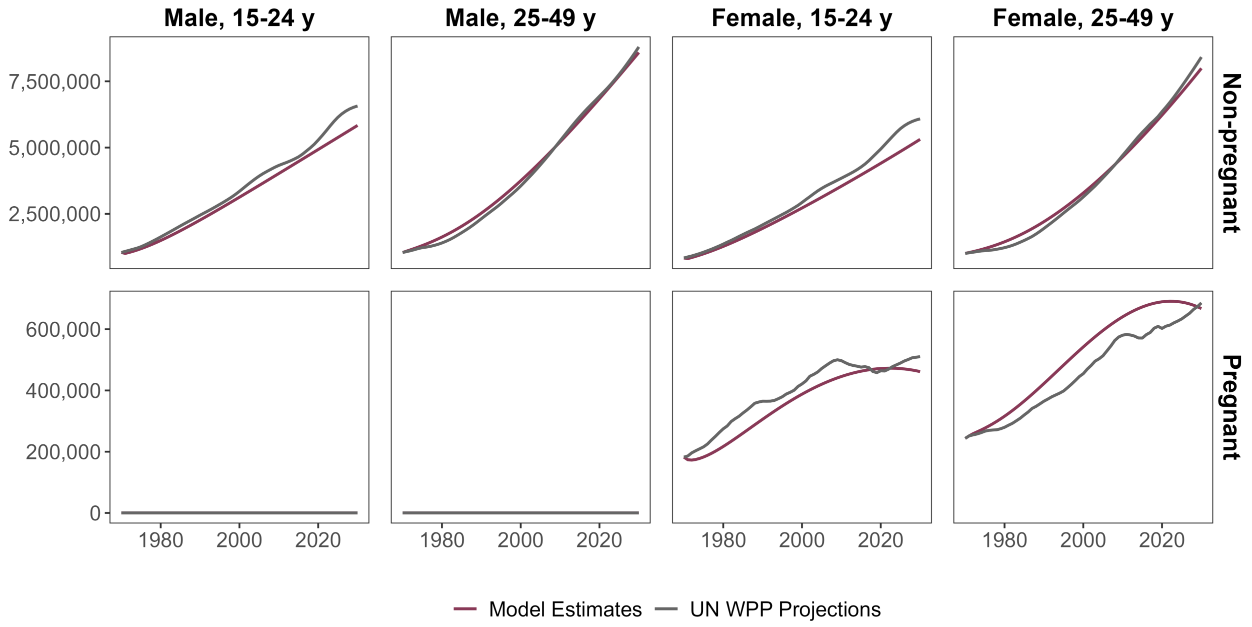

**Figure S4: Comparison of model population estimates with UN WPP projections**

Deviations between model estimates and UN WPP annual projections resulted from demographic parameters in the model, which were derived from cohort-based calculations and linearly interpolated between 1970 and 2030.

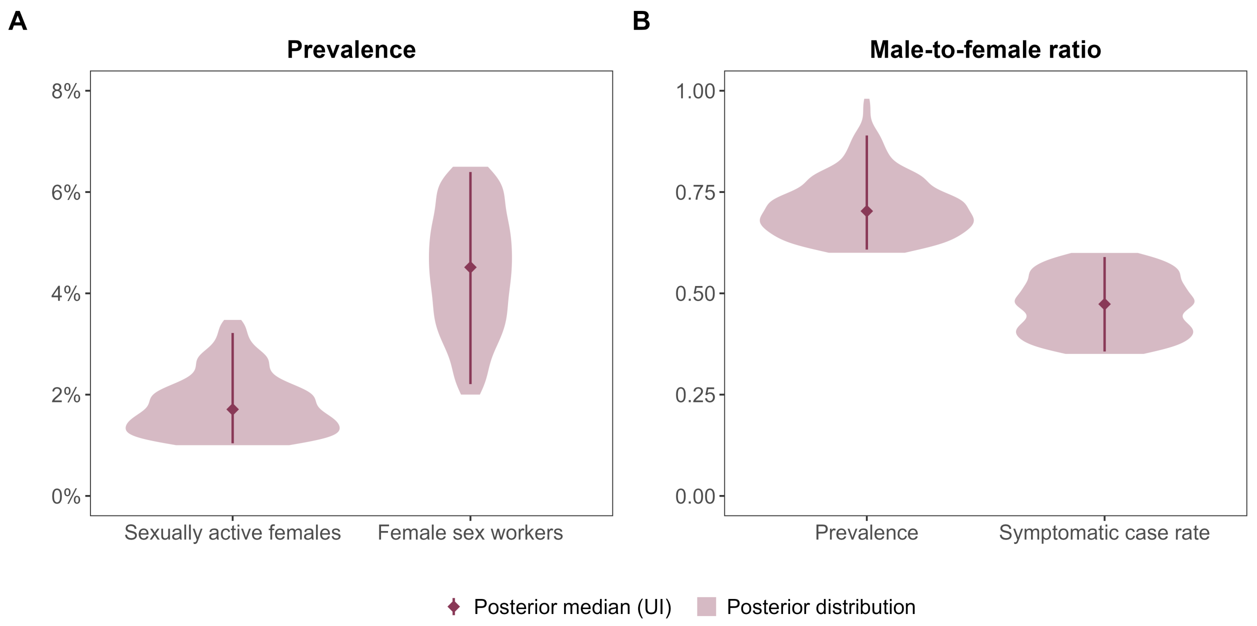

**Figure S5: Distribution of baseline model outcomes after calibration**

Posterior distributions of baseline model outcomes within calibration target ranges at equilibrium in 2025, for (A) gonorrhoea prevalence among sexually active females and FSW and (B) male-to-female ratios for prevalence and symptomatic case rates. Shaded areas represent the distribution of each outcome across retained posterior samples; points show the median, and vertical lines indicate the 95% uncertainty interval.

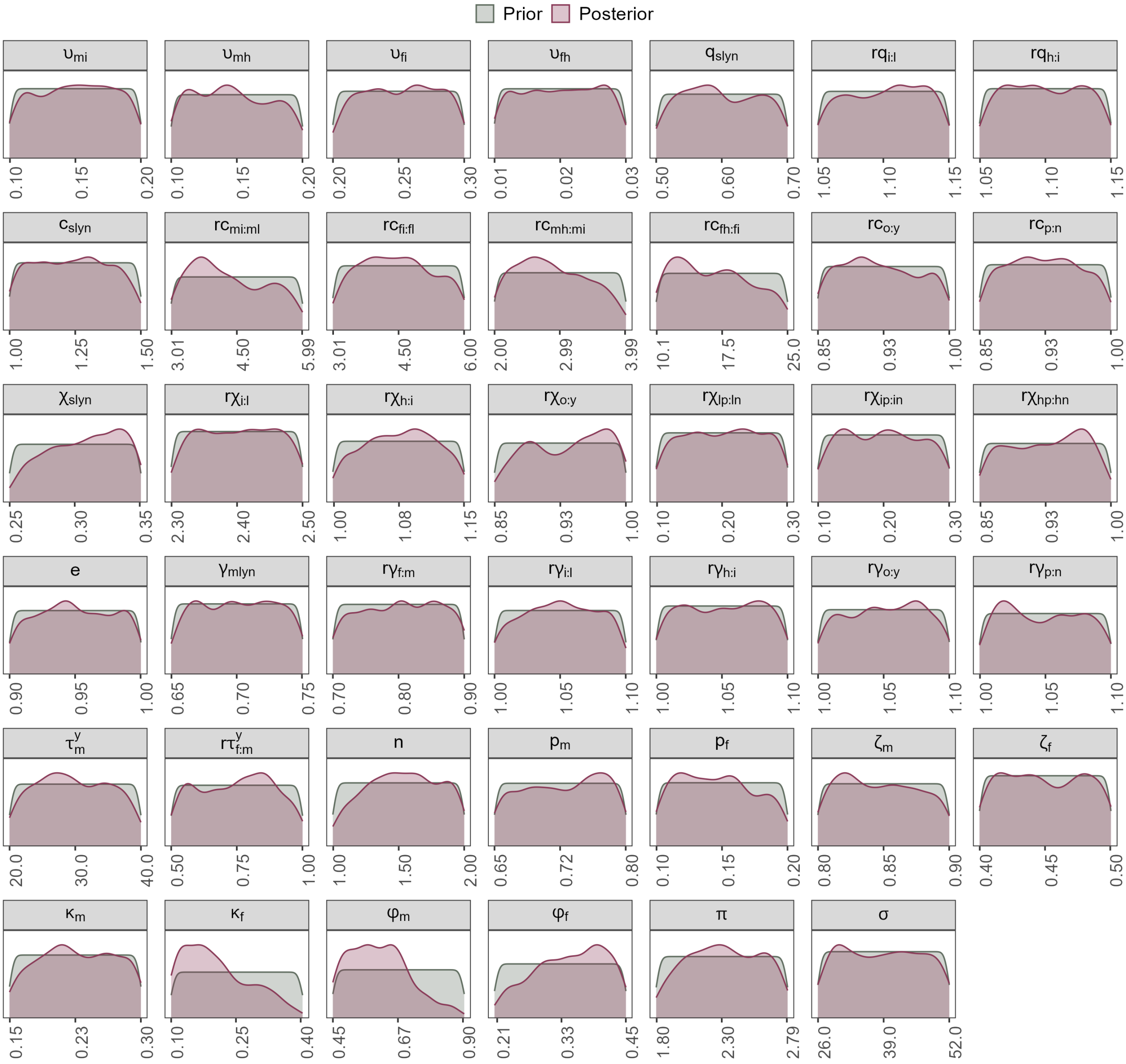

**Figure S6: Prior and posterior distributions for calibrated parameters**

Distributions for calibrated model parameters, stratified by prior (50 000 initial samples) and posterior (536 retained samples). Parameters are defined in Table S2.

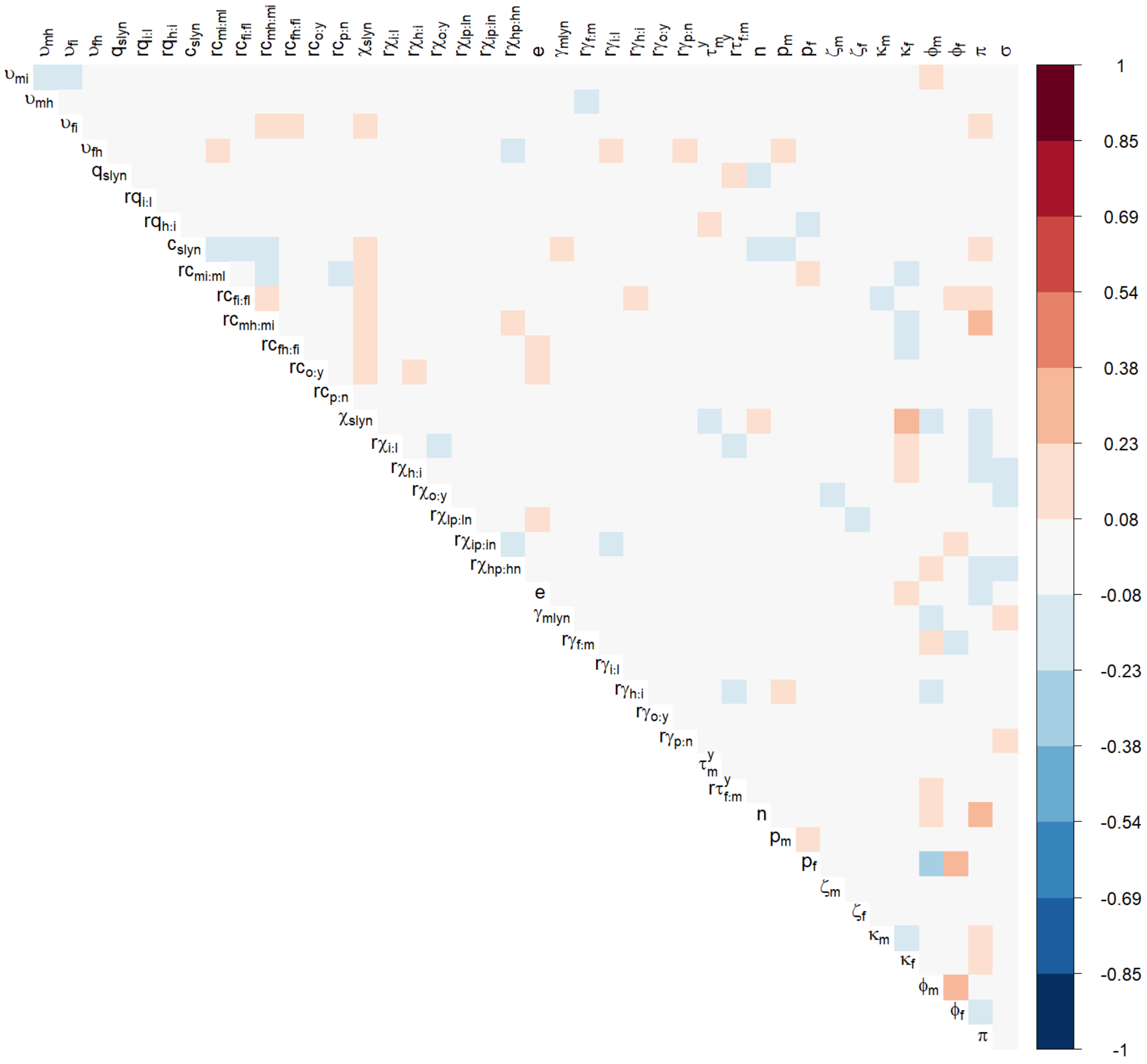

**Figure S7: Rank correlations among posterior samples for calibrated model parameters**

Parameters are defined in Table S2.

#### 2.2 Baseline outcomes

**Table S8: Predicted gonorrhoea burden at baseline, 2025**

| Variable | CFSW | FSW | Men | Pregnant | AGYW |
|---|---|---|---|---|---|
| Population size (millions) | 1.91 (1.35 - 2.58) | 0.26 (0.14 - 0.39) | 13.1 (Fixed)* | 1.16 (1.13 - 1.19) | 4.85 (4.82 - 4.88) |
| Prevalence (%) | 3.56 (1.99 - 6.74) | 4.74 (2.42 - 6.56) | 1.28 (0.74 - 2.34) | 1.89 (1.14 - 3.59) | 1.79 (1.10 - 3.32) |
| Incident infections per 100 individuals, per priority population | 13.1 (6.88 - 24.8) | 11.9 (5.67 - 18.6) | 4.62 (2.70 - 8.29) | 4.97 (2.79 - 9.14) | 4.68 (2.76 - 8.52) |
| QALYs lost per 1000 individuals, per priority population | 0.56 (0.27 - 1.19) | 22.8 (11.1 - 36.0) | 0.21 (0.11 - 0.45) | 80.1 (45.1 - 145) | 1.98 (1.21 - 3.67) |
| Proportion total incident infections in priority population (%) | 21.4 (13.7 - 29.3) | 2.48 (1.00 - 5.55) | 50.0 (45.4 - 55.0) | 5.37 (4.57 - 6.41) | 14.5 (12.6 - 16.5) |
| Proportion total QALYs lost in priority population (%) | 0.84 (0.48 - 1.60) | 4.42 (1.67 - 10.4) | 2.07 (1.31 - 3.58) | 80.6 (76.1 - 83.8) | 5.69 (4.53 - 7.20) |

Results presented as medians with uncertainty intervals and rounded to three significant figures. *Total population size by sex and age is fixed to match annual UN WPP projections.[2] Variation introduced in population size estimate for AGYW due to calibrated probability of being ever sexually active. AGYW: sexually active adolescent girls and young women; CFSW: clients of female sex workers; FSW: female sex workers.

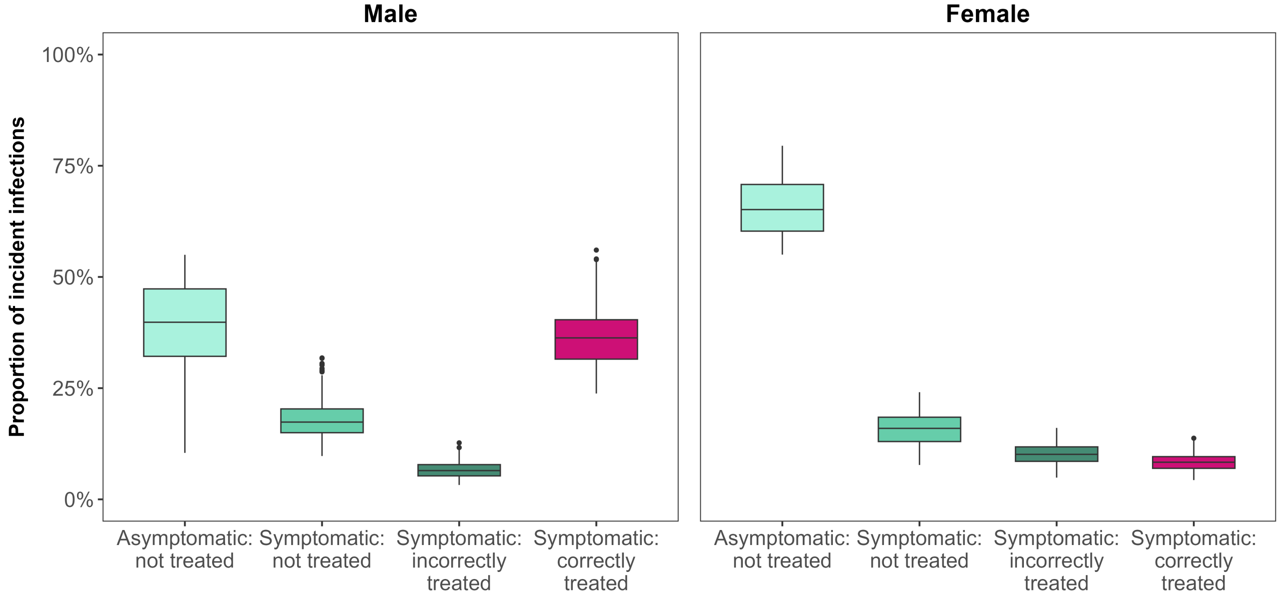

**Figure S8: Distribution of incident gonorrhoea infections by symptom and treatment status at baseline, 2025**

Proportion of incident gonorrhoea infections in 2025 that were asymptomatic and did not access care, symptomatic and did not access care, symptomatic and accessed care but received incorrect treatment, or symptomatic and accessed care and received correct treatment. Proportions presented as medians with 95% uncertainty intervals.

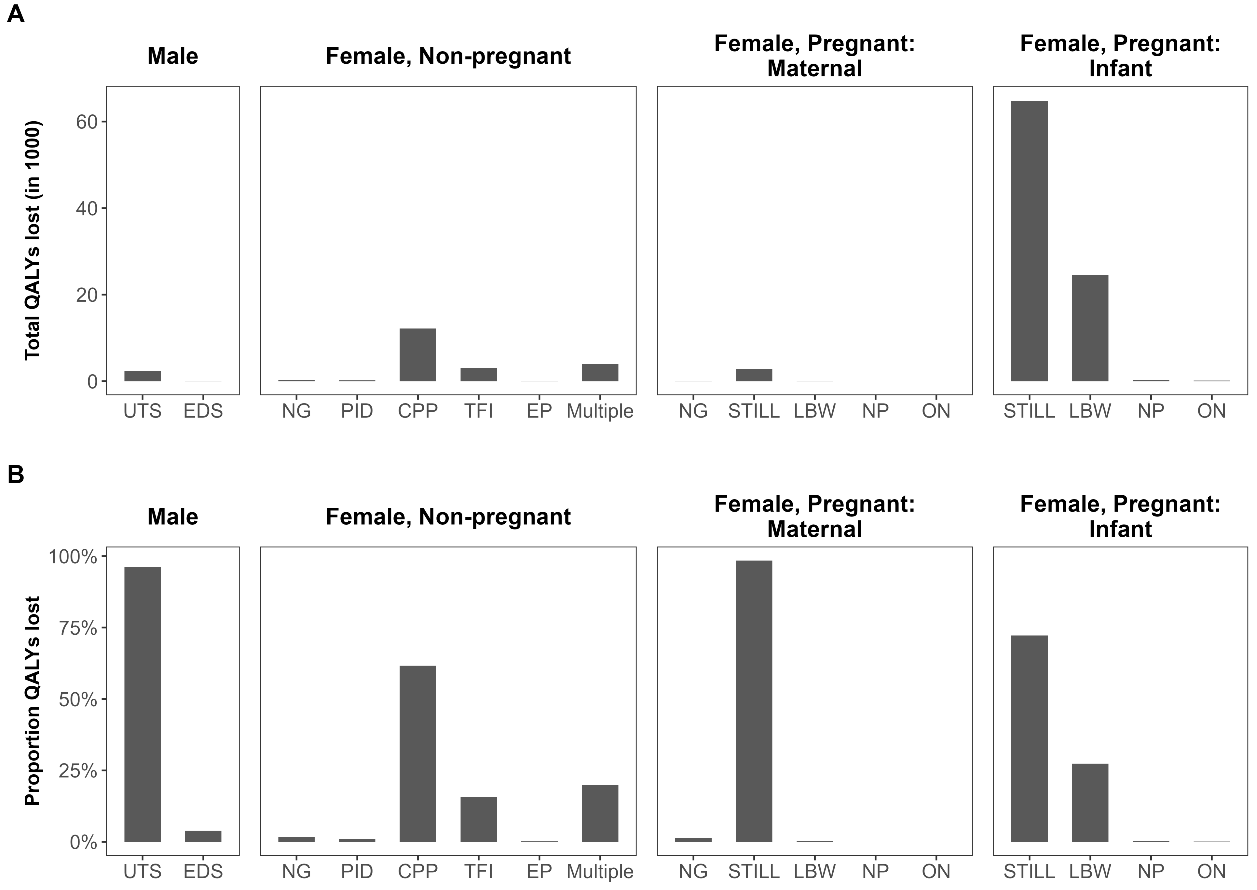

**Figure S9: Decomposition of total discounted lifetime QALYs lost**

Decomposition of total discounted lifetime QALYs lost per population group as (A) number of QALYs lost and (B) proportional distribution of QALYs lost among each population group. Bars represent median estimates. CPP: chronic pelvic pain, EDS: epididymo-orchitis, EP: ectopic pregnancy, LB: live birth, LBW: low birth weight, NG: *Neisseria gonorrhoeae*, NP: neonatal pneumonia, ON: ophthalmia neonatorum, PID: pelvic inflammatory disease, TFI: tubal factor infertility, UTS: urethritis, Multiple: combined sequelae of CPP, TFI, and EP.

#### 2.3 Intervention outcomes

**Table S9: Predicted population-level impact of diagnostic confirmation testing and screening strategies among each priority population with constrained POCT availability, 2025–2030**

| Variable | CFSW | FSW | Men | Pregnant | AGYW |
|---|---|---|---|---|---|
| **Diagnostic confirmation strategy** | | | | | |
| Coverage of POCT among eligible cases* (%) | 18.4 (8.55 - 40.2) | 66.8 (26.0 - 100) | 7.95 (3.91 - 16.2) | 33.6 (15.8 - 71.4) | 13.1 (5.99 - 28.6) |
| Percentage of infections averted relative to baseline (%) | 2.21 (0.77 - 5.31) | 2.07 (0.63 - 5.59) | 1.28 (0.46 - 3.03) | 1.00 (0.47 - 2.12) | 0.93 (0.43 - 1.99) |
| Percentage of QALYs gained relative to baseline (%) | 2.23 (0.78 - 5.28) | 2.17 (0.71 - 5.58) | 1.29 (0.47 - 3.05) | 3.51 (1.75 - 7.22) | 0.85 (0.39 - 1.82) |
| Infections averted per 100 tests | 99.8 (45.7 - 165) | 94.3 (41.9 - 193) | 58.7 (27.1 - 96.3) | 44.1 (28.1 - 66.2) | 40.6 (26.3 - 64.0) |
| QALYs gained per 100 tests | 10.4 (4.83 - 17.8) | 10.1 (5.33 - 20.0) | 6.07 (2.90 - 10.1) | 16.2 (11.2 - 23.1) | 3.80 (2.54 - 5.82) |
| **Screening strategy** | | | | | |
| Coverage of POCT among eligible cases* (%) | 0.98 (0.71 - 1.41) | 5.77 (3.90 - 11.1) | 0.14 (0.14 - 0.15) | 0.82 (0.80 - 0.85) | 0.52 (0.47 - 0.60) |
| Percentage of infections averted relative to baseline (%) | 0.58 (0.41 - 0.82) | 0.62 (0.20 - 1.52) | 0.12 (0.10 - 0.13) | 0.08 (0.06 - 0.10) | 0.13 (0.11 - 0.16) |
| Percentage of QALYs gained relative to baseline (%) | 0.58 (0.41 - 0.81) | 0.65 (0.24 - 1.50) | 0.11 (0.10 - 0.13) | 0.30 (0.25 - 0.34) | 0.12 (0.10 - 0.14) |
| Infections averted per 100 tests | 25.5 (14.6 - 44.2) | 26.9 (9.97 - 59.1) | 5.04 (3.15 - 8.16) | 3.51 (2.04 - 5.97) | 5.89 (3.56 - 9.87) |
| QALYs gained per 100 tests | 2.64 (1.44 - 4.70) | 2.89 (1.20 - 6.04) | 0.52 (0.31 - 0.87) | 1.32 (0.80 - 2.42) | 0.55 (0.34 - 0.92) |

Results presented as medians with uncertainty intervals and rounded to three significant figures. Constrained POCT availability was modelled as 25 000 tests annually. *Cases refer to the number of individuals accessing care for vaginal or urethral discharge (diagnostic scenario) or attending routine healthcare services (screening scenario). Individuals may be counted more than once due to repeat infections or multiple healthcare visits. AGYW: sexually active adolescent girls and young women; CFSW: clients of female sex workers; FSW: female sex workers.

**Table S10: Predicted population-level impact of diagnostic confirmation testing and screening strategies among each priority population with unrestricted POCT availability, 2025–2030**

| Variable | CFSW | FSW | Men | Pregnant | AGYW |
|---|---|---|---|---|---|
| **Diagnostic confirmation strategy** | | | | | |
| Average number POCT used annually (×100 000) | 1.39 (0.65 - 2.97) | 0.38 (0.13 - 0.99) | 3.19 (1.59 - 6.46) | 0.72 (0.35 - 1.54) | 1.96 (0.90 - 4.28) |
| Percentage of infections averted relative to baseline (%) | 11.6 (4.87 - 24.1) | 3.07 (0.73 - 10.0) | 15.3 (6.81 - 28.9) | 3.05 (1.64 - 4.97) | 6.93 (3.81 - 10.7) |
| Percentage of QALYs gained relative to baseline (%) | 11.6 (4.87 - 24.1) | 3.33 (0.83 - 10.1) | 15.4 (6.86 - 28.9) | 10.7 (6.43 - 15.8) | 6.28 (3.45 - 9.76) |
| Infections averted per 100 tests | 102.0 (46.2 - 171) | 93.9 (41.9 - 191) | 60.3 (27.4 - 99.8) | 43.9 (28.1 - 66.0) | 39.7 (25.9 - 62.1) |
| QALYs gained per 100 tests | 10.5 (4.83 - 18.2) | 10.1 (5.32 - 20.0) | 6.15 (2.92 - 10.5) | 16.1 (11.1 - 22.8) | 3.73 (2.49 - 5.70) |
| **Screening strategy** | | | | | |
| Average number POCT used annually (×100 000) | 25.4 (18.0 - 34.4) | 4.33 (2.25 - 6.38) | 175.4 (175.1 - 175.6) | 30.4 (29.7 - 31.1) | 47.6 (42.8 - 52.1) |
| Percentage of infections averted relative to baseline (%) | 36.3 (27.7 - 42.2) | 7.25 (2.16 - 19.5) | 45.8 (40.3 - 49.1) | 7.59 (6.07 - 9.53) | 17.46 (14.6 - 20.5) |
| Percentage of QALYs gained relative to baseline (%) | 36.1 (27.9 - 41.9) | 7.65 (2.50 - 19.7) | 45.5 (40.1 - 48.5) | 28.0 (24.6 - 31.8) | 15.9 (13.3 - 18.8) |
| Infections averted per 100 tests | 15.6 (8.48 - 28.6) | 19.3 (6.96 - 42.0) | 2.84 (1.68 - 4.86) | 2.76 (1.60 - 4.73) | 4.06 (2.38 - 6.88) |
| QALYs gained per 100 tests | 1.60 (0.85 - 2.95) | 2.08 (0.86 - 4.31) | 0.29 (0.17 - 0.52) | 1.03 (0.62 - 1.90) | 0.38 (0.23 - 0.66) |

Results presented as medians with uncertainty intervals and rounded to three significant figures. Unrestricted POCT availability was modelled as all eligible cases tested. Cases refer to the number of individuals accessing care for vaginal or urethral discharge (diagnostic scenario) or attending routine healthcare services (screening scenario). Individuals may be counted more than once due to repeat infections or multiple healthcare visits. AGYW: sexually active adolescent girls and young women; CFSW: clients of female sex workers; FSW: female sex workers.

#### 2.4 Sensitivity analyses

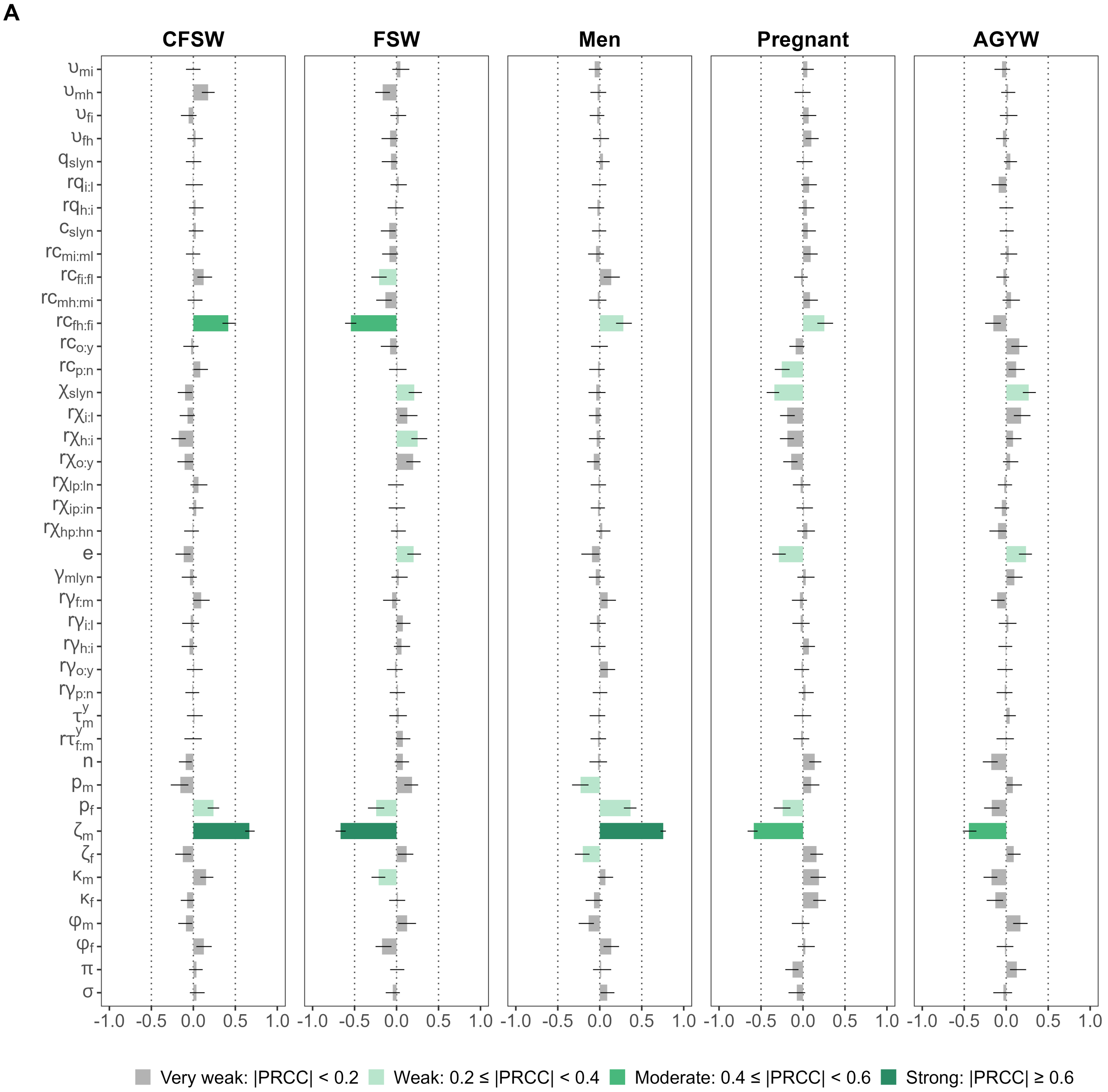

**Figure S10: Correlation between parameter values and priority population prioritisation order by test strategy**

Partial rank correlation coefficients between posterior parameter values and the prioritisation order of each priority population according to the proportion of incident gonorrhoea infections averted under constrained POCT availability (25 000 annually) between 2025 and 2030 for (A) diagnostic confirmation and (B) screening strategies across each priority population. Bars and lines indicate median and 95% confidence intervals. Parameters are defined in Table S2.

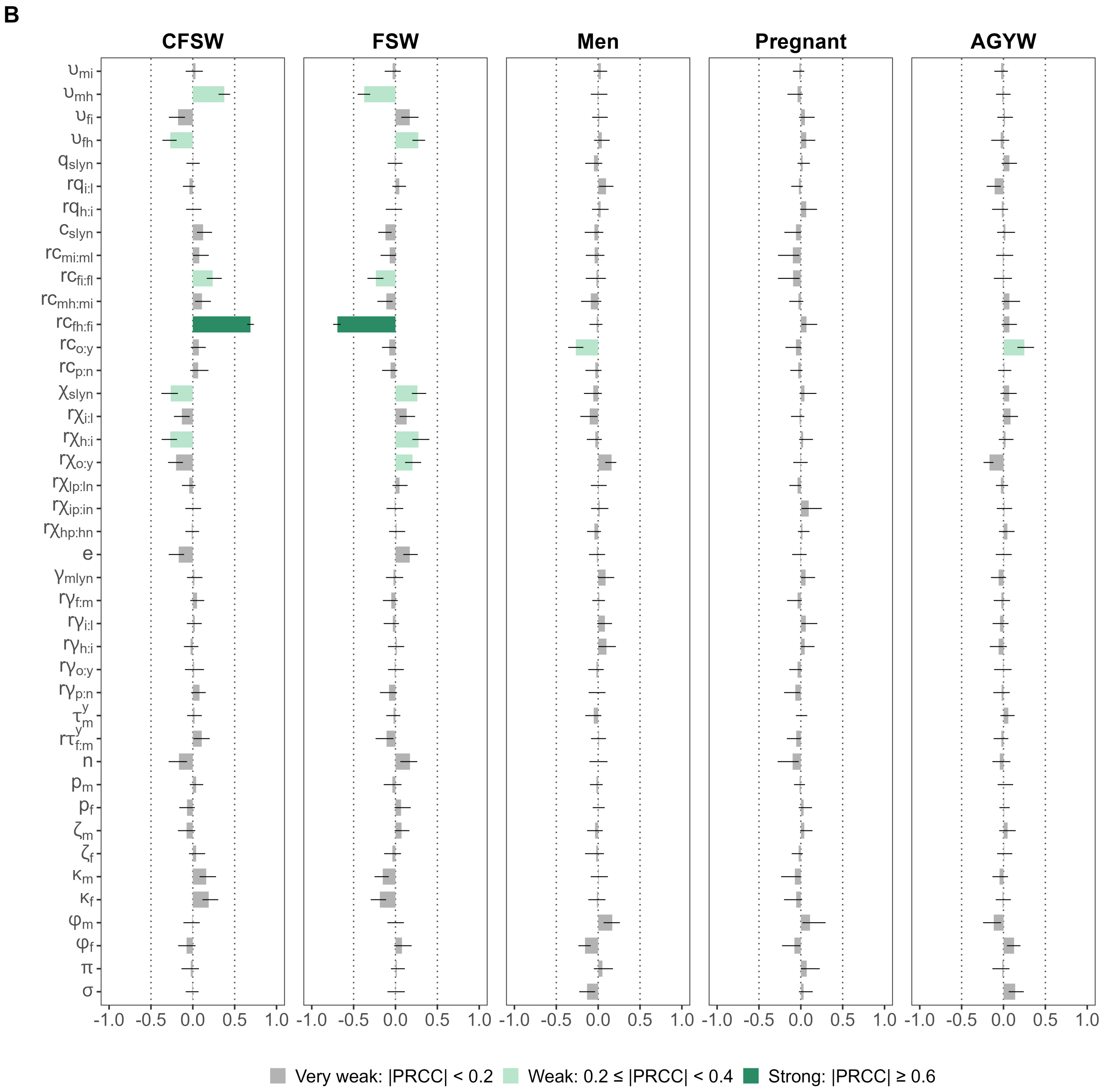

**Figure S10: Correlation between parameter values and priority population prioritisation order by test strategy (continued)**

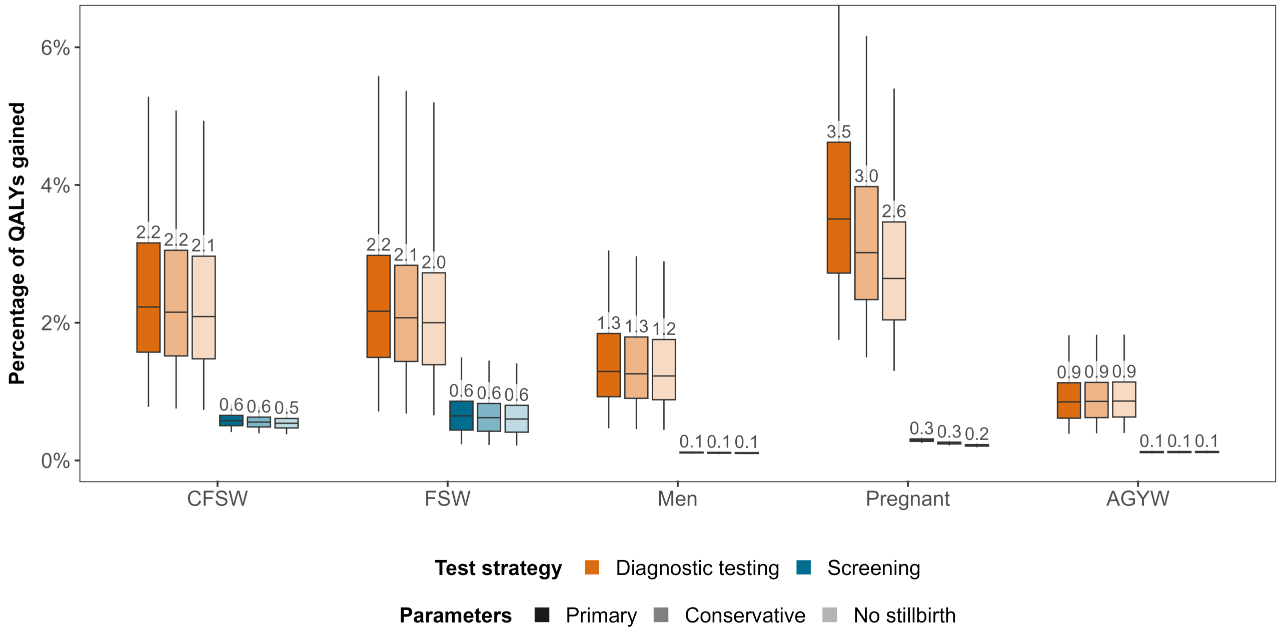

**Figure S11: Influence of parameter assumptions for pregnancy-related sequelae on the predicted population-level impact of diagnostic confirmation and screening strategies, 2025–2030**

Population-level impact of deploying gonorrhoea POCTs (25 000 annually) among each of five priority populations between 2025 and 2030 through either diagnostic confirmation among individuals accessing care for urethral or vaginal discharge, or screening of attendees at routine healthcare services. Impact was assessed as the percentage of lifetime QALYs gained relative to the baseline scenario of syndromic management only, under three sets of parameter assumptions: (i) Primary, as used in the main analysis, (ii) Conservative, which increased the utility weight for stillbirth from 0 to 0.5, and reduced the duration of infant LBW-associated losses from lifetime to 10 years, and (iii) No stillbirth, which increased the utility weight for stillbirth from 0 to 1 to exclude stillbirth QALY losses for infants. Parameters describing maternal QALY losses for stillbirth and LBW remained unchanged. Boxplots represent the median (horizontal line), interquartile range (box), and uncertainty interval (whiskers). AGYW: sexually active adolescent girls and young women; CFSW: clients of female sex workers; FSW: female sex workers; POCT: point-of-care test; LBW: low birth weight; QALY: quality-adjusted life year.

3. Kenya National Bureau of Statistics, Kenya Ministry of Health, ORC Macro. Kenya Demographic and Health Survey 2003. Calverton, Maryland: CBS, MOH, and ORC Macro; 2004.

4. Kenya National Bureau of Statistics and ICF Macro. Kenya Demographic and Health Survey 2008-09. Calverton, Maryland: KNBS and ICF Macro; 2010.

5. Kenya National Bureau of Statistics. Kenya Demographic and Health Survey 2014. Rockville, Maryland: KNBS and ICF; 2015.

6. KNBS and ICF. Kenya Demographic and Health Survey 2022. Nairobi, Kenya and Rockville, Maryland, USA: KNBS and ICF; 2023.

7. National AIDS and STI Control Programme. Kenya Population-based HIV Impact Assessment. Nairobi: NASCOP; 2022.

20. NASCOP. National behavioral assessment of key populations in Kenya. Polling booth survey report; 2014.

21. NASCOP. National behavioural assessment of key populations in Kenya. Polling booth survey report; 2016.

22. NASCOP. Third national behavioural assessment of key populations in Kenya. Polling booth survey report; 2018.

43. Kenya institute for public policy research and analysis. An assessment of healthcare delivery in Kenya under the devolved system. Special paper no. 19; 2018.

44. Kenya Ministry of Health. Kenya health facility census report. Nairobi: Kenya Ministry of Health; 2023.

45. Otieno PO, Kiroro F, Runyenje C, Kamau P. Unmet need for primary healthcare and associated individual and household-level factors in Kenya: results from a national survey. BMJ Open. 2021;11:41032.

46. Parmley L, Rao A, Kose Z, Lambert A, Max R, Phaswana-Mafuya N, et al. Antenatal care presentation and engagement in the context of sex work: Exploring barriers to care for sex worker mothers in South Africa. Reproductive Health. 2019 5;16(1):1-12.

47. Moore BE, Govaerts L, Kapadia F. Maternal health and maternal health service utilization among female sex workers: A scoping review. Women's Health. 2023 1;19.

48. World Health Organization. Guidelines for the management of symptomatic sexually transmitted infections. Web Annex A. Syndromic management or point of care tests for urethral discharge: systematic review and mathematical modelling. Geneva: World Health Organization; 2021.

57. Wiesner PJ, Thompson SE. Gonococcal diseases. Year Book Medical Publishers; 1980.

63. World Health Organization. Guidelines for the management of symptomatic sexually transmitted infections. Geneva: World Health Organization; 2021.

92. Institute for Health Metrics and Evaluation. Global Burden of Disease Study 2019 (GBD 2019) Disability Weights; 2020.

93. World Health Organization. The global burden of disease: 2004 update. Geneva: World Health Organization; 2008.

94. Institute of Medicine (US) Committee to Study Priorities for Vaccine Development. Vaccines for the 21st Century: A Tool for Decisionmaking. Stratton KR, Durch JS, Lawrence RS, editors. Washington: National Academies Press (US); 2000.

95. Aledort JE, Hook EW, Weinstein MC, Goldie SJ. The cost effectiveness of gonorrhea screening in urban emergency departments. Sexually transmitted diseases. 2005 7;32(7):425-36.

96. Jamison DT, Shahid-Salles SA, Jamison J, Lawn JE, Zupan J. Incorporating Deaths Near the Time of Birth into Estimates of the Global Burden of Disease. Lopez A, Mathers C, Ezzati M, Jamison D, Murray CJL, editors. New York: Oxford University Press; 2006.

97. Camacho EM, Gold KJ, Murphy M, Storey C, Heazell AEP. Measuring EQ-5D-5L utility values in parents who have experienced perinatal death. The European Journal of Health Economics. 2024 11;25(8).

98. Lee K, You S, Li Y, Chesson H, Gift TL, Berruti AA, et al. Estimation of the Lifetime Quality-Adjusted Life Years (QALYs) Lost Due to Syphilis Acquired in the United States in 2018. Clinical Infectious Diseases. 2023 2;76(3):E810-9.

99. Ong JJ, Chen M, Hocking J, Fairley CK, Carter R, Bulfone L, et al. Chlamydia screening for pregnant women aged 16-25 years attending an antenatal service: a cost-effectiveness study. BJOG. 2016 6;123(7):1194-202.

100. Trikalinos T. Technical Appendix: Probability of pelvic inflammatory disease in Chlamydia trachomatis infection; 2018.

106. Institute of Medicine. Preterm birth: Causes, consequences, and prevention. Behrman RE, Butler AS, editors. Washington, DC: National Academies Press; 2007.

107. van der Pal S, Steinhof M, Grevinga M, Wolke D, Verrips GE. Quality of life of adults born very preterm or very low birth weight: A systematic review. Acta paediatrica. 2020 10;109(10):1974-88.

108. Murray CJL, Ezzati M, Flaxman AD, Lim S, Lozano R, Michaud C, et al. GBD 2010: design, definitions, and metrics. The Lancet. 2012 12;380(9859):2063-6.

109. World Health Organization Department of Data and Analytics. WHO methods and data sources for global burden of disease estimates 2000-2019. Geneva: World Health Organization; 2020.
